# Patterns of healthcare use in people with narcolepsy: a population-based cohort study in England

**DOI:** 10.64898/2026.07.23.26358663

**Authors:** Helen Strongman, Aurélien Belot, Hema Mistry, Ellen Nolte, Sofia H Eriksson, Michelle A Miller, Ian E Smith, Charlotte Warren-Gash, Krishnan Bhaskaran

## Abstract

People with narcolepsy experience delays in diagnosis and inconsistent post-diagnosis care, but the pattern and scale of their healthcare use is poorly described. In this population-based cohort study, we used primary care and linked hospital activity data to compare healthcare use in people with narcolepsy (n=2,772) and a matched comparison group in England (n=13,860). Narcolepsy was defined by a first coded record in primary care or admitted care data between 02 January 1998 and 31 December 2019; this date was the index date for both groups. People with narcolepsy had approximately double the rate of healthcare use in the period from five years before to five years after the index date. Annually, this corresponded on average to an additional 1.9 (95% confidence interval (CI) 1.8-2.0) outpatient events, 0.36 (95% CI 0.30-0.42) admitted patient care events, 0.25 (95% CI 0.22-0.29) Accident & Emergency events and 4.3 (95% CI 3.9-4.7) primary care events per person. Service use in all settings peaked at index and remained elevated for at least 15 years either side. The elevated rates of possible-sleep related outpatient events (respiratory, neurology paediatric, and ear nose & throat) peaked in the year including and after the index date, at 1.50 (95% CI 1.41-1.59); before declining to <0.5 visits per person-year after five years. Our findings of sustained elevated use of healthcare by people with narcolepsy across NHS settings may reflect diagnostic delay, comorbidities, and ongoing narcolepsy-related healthcare needs being met largely outside specialist sleep care, highlighting opportunities to improve healthcare services.

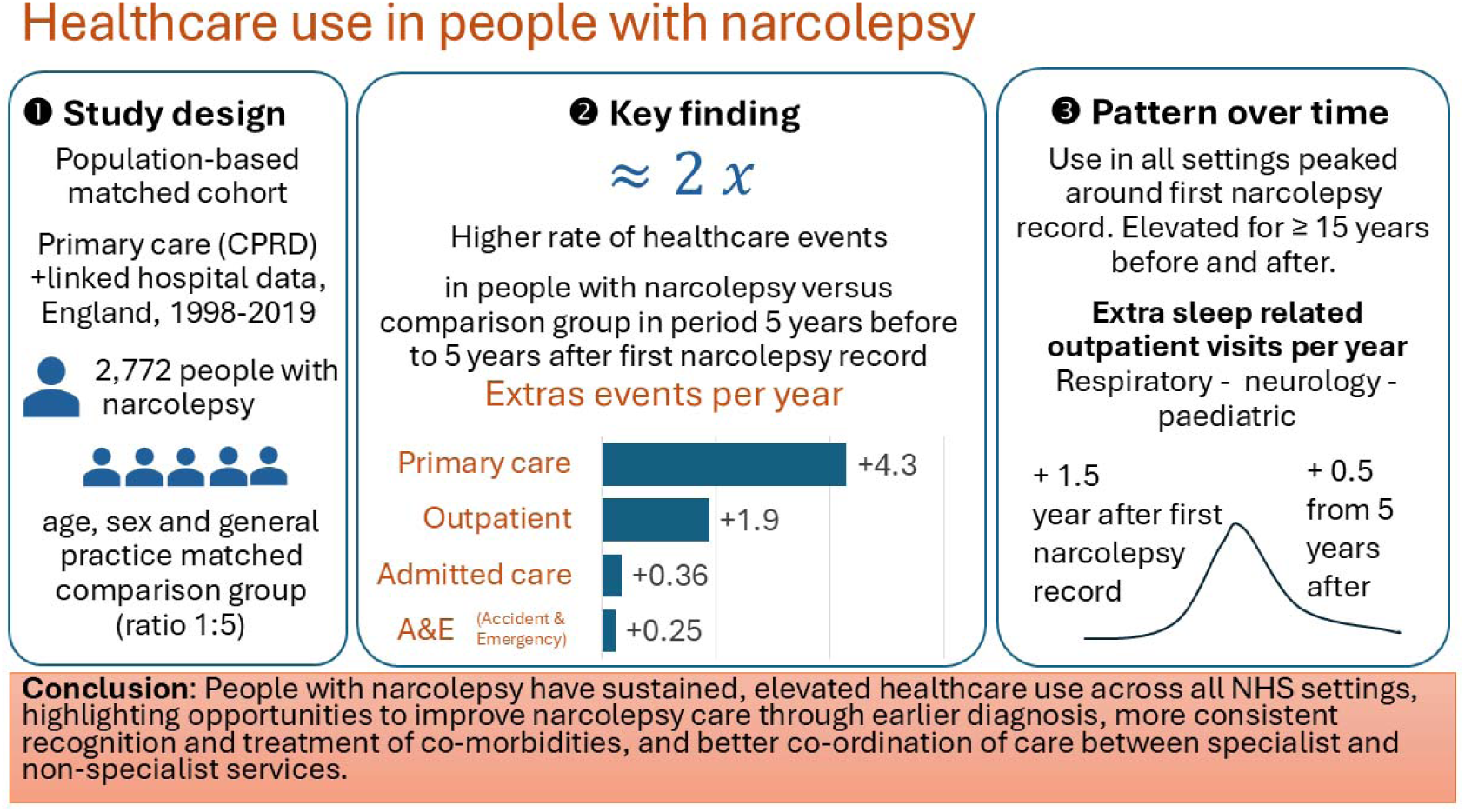
GRAPHICAL ABSTRACT.

## INTRODUCTION

Narcolepsy is a chronic neurological condition that affects approximately 1 in 2,000 people[1] and limits participation in education, work, family and social activities[2,3]. Symptoms include excessive daytime sleepiness, disturbed nighttime sleep, and in people with type 1 narcolepsy, cataplexy[3]. In the UK, people with narcolepsy symptoms mostly present to general practitioners (GPs) before they are referred to the appropriate specialist service for assessment. This is usually a neurology, respiratory or paediatric-led sleep medicine centre with facilities for objective testing, including overnight polysomnography[4]. Narcolepsy treatment includes medications prescribed directly by sleep specialists or through coordinated care with GPs. Non-pharmaceutical treatments such as scheduled naps[3,5] are also recommended. Treatment should be reviewed regularly with specialist oversight, including monitoring of effectiveness, side effects, and co-morbidities[6–8].

Optimising the diagnosis and treatment pathways of people with narcolepsy requires good understanding of how they currently use healthcare. Research from Europe, North America and Asia suggests that people with narcolepsy have a higher use of secondary care services than those without [9–16]. Studies found that, compared to matched controls, people with narcolepsy have higher rates of outpatient visits[9–11,15,16], inpatient admissions [9–12,14,15], emergency department visits[10,12,14,16], neurology visits[14,16], psychiatry visits[14,16] and respiratory visits[14]. Jennum et al. documented that healthcare use in this patient group increased in the years before diagnosis, peaked at diagnosis, and remained elevated for at least 11 years thereafter [9].

However, important knowledge gaps remain. First, primary care, the most common point of contact in many health systems including the National Health Service (NHS) in England[17], has received little attention. Secondly, published studies have tended to focus on specialist service use overall without distinguishing the range of specialties providing healthcare services. Third, we identified only two studies that explored patterns of healthcare use among people before and after diagnosis; neither of these was set in England[9,18].

This study aimed to contribute to closing this important evidence gap by estimating and comparing rates of healthcare use in people with narcolepsy and a matched comparison group in all NHS settings in England and exploring how rates in each setting change over time before and after diagnosis.

## METHODS

### Study design and setting

In this population-based cohort study, we analysed Clinical Practice Research Datalink (CPRD) primary care records[19,20] linked to external data sets[21] including Hospital Episode Statistics (HES) Admitted Patient Care (APC)[22], HES Outpatient, HES Accident and Emergency (A&E), Office of National Statistics (ONS) mortality [23], GP practice level small area-based deprivation, and patient level small-area based urban rural data[24]. Primary care records are collected by NHS GP practices to support practice administration and clinical care; HES datasets underpin service management, commissioning, and reimbursement within the NHS through collection of national hospital activity data.

Digital object identifiers (DOIs) and further documentation describing each data source and associated data governance arrangements are listed in Supplementary Appendix table 1. Primary care data were drawn from both CPRD databases, CPRD Aurum and CPRD GOLD, which include practices using different clinical software systems. In combination, the versions of CPRD data used for this analysis represent approximately one quarter of the UK population. Linked data availability, and therefore this study, is restricted to general practices in England.

**Table 1:** Characteristics of narcolepsy and comparison group.

|  | **Narcolepsy group** | **Comparison group** |
| --- | --- | --- |
| **N+** | 2,772 | 13,860 |
| **Follow-up time, median (IQR) person-years** |  |  |
| Total | 13.6 (7.2, 17.9) | 14.5 (7.7, 18.8) |
| Before index date | 5.6 (2.0, 10.8) | 5.9 (2.4, 11.1) |
| After index date | 4.8 (2.2, 9.8) | 5.4 (2.4, 10.3) |
| Primary analysis* | 7.6 (5.6, 10.0) | 8.0 (5.9, 10.0) |
| Total (HES Outpatient) | 10.9 (5.5, 15.8) | 11.6 (6.1, 16.3) |
| Total (HES A&E) | 10.8 (5.3, 15.5) | 11.4 (5.9, 16.0) |
| **Age at index (years)** |  |  |
| Mean (SD) | 42.2 (20.3) | 43.2 (20.5) |
| Median (IQR) | 39.0 (25.0, 55.0) | 39.0 (25.0, 55.0) |
| **Sex** |  |  |
| male | 1,309 (47.2) | 6,545 (47.2) |
| female | 1,463 (52.8) | 7,315 (52.8) |
| **Calendar year at index** |  |  |
| 1998-1999 | 131 (4.7) | 655 (4.7) |
| 2000-2004 | 569 (20.5) | 2,845 (20.5) |
| 2005-2009 | 616 (22.2) | 3,080 (22.2) |
| 2010-2014 | 697 (25.1) | 3,485 (25.1) |
| 2015-2019 | 759 (27.4) | 3,795 (27.4) |
| **Ethnicity** |  |  |
| White | 2,353 (84.9) [87.2] | 10,743 (77.5) [86.2] |
| South Asian | 74 (2.7) [2.8] | 699 (5.0) [5.6] |
| Black | 172 (6.2) [6.4] | 543 (3.9) [4.4] |
| Other | 35 (1.3) [1.3] | 280 (2.0) [2.2] |
| Mixed | 48 (1.7) [1.8] | 197 (1.4) [1.6] |
| missing | 90 (3.2) | 1,398 (10.1) |
| **Area-based deprivation quintile (Carstairs)** |  |  |
| 1 (least deprived) | 306 (11.0) | 1,530 (11.0) |
| 2 | 504 (18.2) | 2,520 (18.2) |
| 3 | 685 (24.7) | 3,425 (24.7) |
| 4 | 607 (21.9) | 3,035 (21.9) |
| 5 (most deprived) | 670 (24.2) | 3,350 (24.2) |
| **Urban Rural** |  |  |
| urban | 2,379 (85.8) | 11,835 (85.4) |
| rural | 390 (14.1) | 1,996 (14.4) |
| missing | 3 (0.1) | 29 (0.2) |
| **Practice size quintile** |  |  |
| 1 largest | 971 (35.0) | 4,855 (35.0) |
| 2 | 670 (24.2) | 3,350 (24.2) |
| 3 | 538 (19.4) | 2,690 (19.4) |
| 4 | 358 (12.9) | 1,790 (12.9) |
| 5 smallest | 235 (8.5) | 1,175 (8.5) |
| **Practice size** |  |  |
| Median (IQR) | 38041 (25361, 55103) | 38041 (25361, 55103) |

Footnote: Categorical figures are presented as number (% of total). \*The primary analysis is censored five years before and after the index date. Numbers in square brackets are percentages excluding missing values. IQR Interquartile range HES Hospital Episode Statistics A&E Accident & Emergency

### Study population

We defined the study population and exposure time as individuals registered with CPRD-contributing practices that passed basic CPRD quality checks, had a recorded sex, were eligible for linkage to HES APC data and alive and registered in the practice for at least 90 days during the period covered by the linked data (2 January 1998 to 29 March 2021). There was no age restriction. In view of substantial changes in primary care access and coding practices during the COVID-19 pandemic[25], our study period ended on 31 December 2019. The study period was therefore 2 January 1998 to 31 December 2019. To prevent duplication of individuals at the same point in time, we removed data from practices contributing to both CPRD GOLD and CPRD Aurum, and CPRD Aurum practices that subsequently merged into other contributing practices. Analyses using HES Outpatient and HES A&E data were restricted to people eligible for linkage to the relevant dataset.

Procedures used to define the matched cohort and estimate outcomes and covariates are detailed in Figure 1.

**Figure 1:**
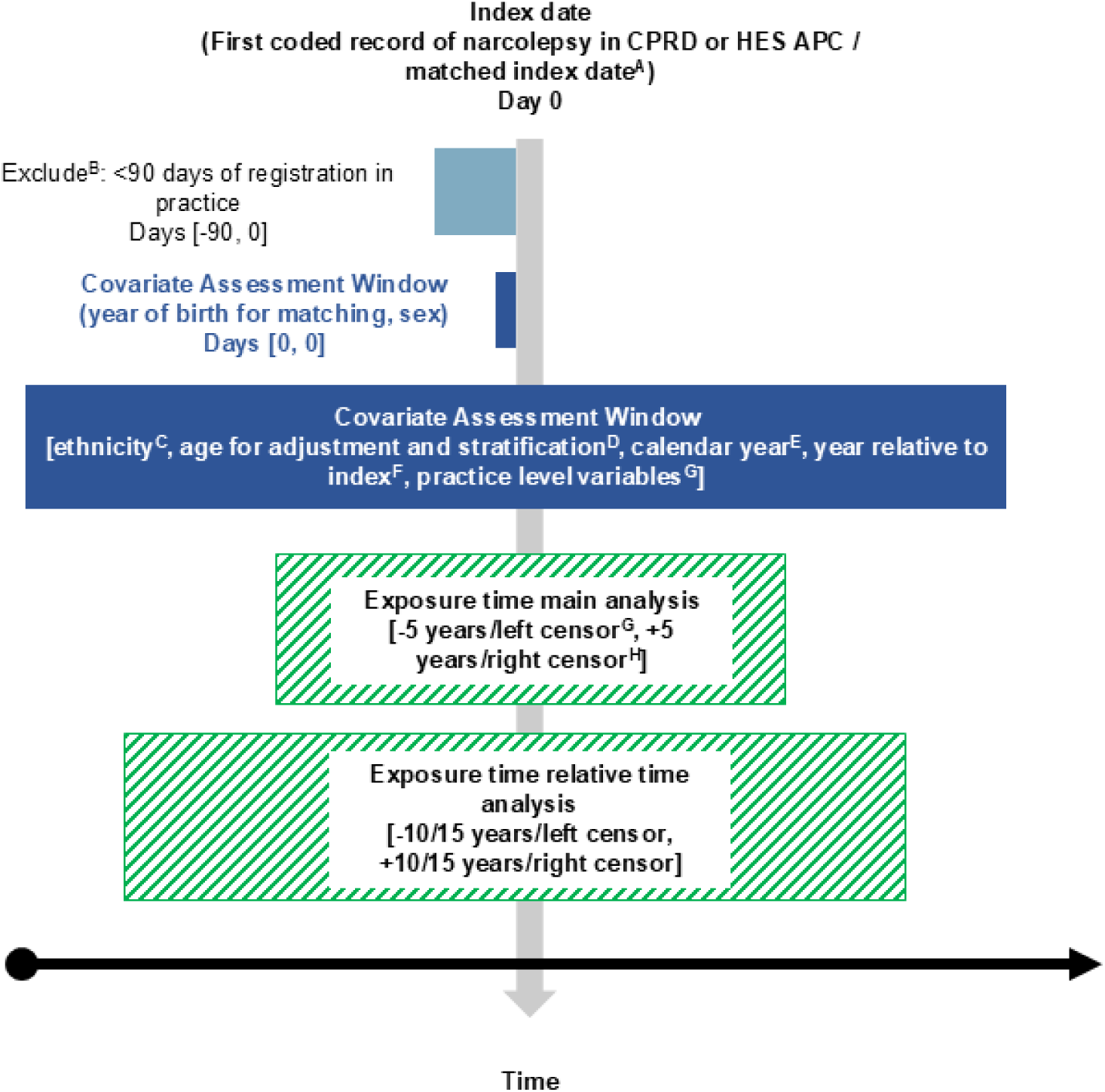
Procedures used to identify the study cohort. A Each person with narcolepsy was matched to up to 5 individuals without narcolepsy by year of birth (plus/minus 3 years with nearest match prioritised), sex, general practice, and registration time whereby the matched person was actively registered in the practice on the index date of the person with narcolepsy and had at least 90 days follow-up prior to this date. The index date for the comparison group matched the index date of the matched individual in the narcolepsy group. People were matched in calendar time order to avoid time-related biases. To represent the full population at risk, individuals with narcolepsy were available to be selected as comparators, up to the index date (incidence density sampling). People were matched in calendar time order to avoid time-related biases; matching was without replacement. B Chronic conditions may be recorded shortly after a patient joins a new practice and misclassified as incident rather than prevalent cases. We therefore required a minimum period of 90 days registration before the index date. C Ethnicity was measured using the most commonly recorded ethnicity in primary care data (or latest if equally common). Unknown and missing values were replaced with the most commonly reported ethnicity in HES data where available. D Age was derived by subtracting the birth day set at 01 July for each individual from the index date. Age was time updated in the analysis dataset (see E) and modelled as a 3-knotted restricted cubic spline in regression models. E Datasets including exposures and covariates for the full cohort were time split by calendar year and year relative to diagnosis. Outcomes datasets for each data source were merged with this time split file: additional rows were created when more than one outcome event occurred within a calendar year/year relative to diagnosis episode. This ensured that events to different specialties on the same day are counted correctly in specialty-specific analyses. In the all events analysis, visits to different outpatient specialists or primary care / consultation type combinations on the same day were counted as unique events, HES A&E visits on the same day and HES APC episodes in the same admission were counted as a single event. F Practice area-based deprivation (Carstairs index) and patient area-based urban rural status were measured using postcodes at the time of CPRD data collection linked to the 2011 census. Practice size was estimated as the CPRD denominator population registered in the practice on 01/07/2019. G Exposure time was left censored at the latest of the ONS mortality coverage start date [02/01/1998] or practice registration date plus 90 days. Analyses using HES Outpatient data and HES Accident & Emergency data were left centred on 01/04/2003 in line with their coverage periods. H Exposure time was right censored at the earliest of end of registration in practice, date of death recorded in Office for National Statistics mortality data or 31/12/2019. In the comparison group, exposure time was further censored at the first record of narcolepsy.

### Matched cohort (exposures)

The exposed group comprised individuals diagnosed with narcolepsy identified using validated codelists and algorithms[26]. The index date was defined as the event date of the first ever coded record of narcolepsy in primary care or HES APC data. We excluded individuals with a narcolepsy code before the start of exposure time or one or more narcolepsy codes that lacked a valid event date.

The comparison group was selected from people in the study population without a diagnosis of narcolepsy. We matched each person with narcolepsy to up to 5 individuals without narcolepsy by year of birth (plus/minus 3 years), sex, GP practice, and registration time. The index date of the matched individual in the narcolepsy group was assigned as the index date for the individual in the comparison group.

### Outcomes

Outcomes included counts of attended outpatient events (HES OP data), admitted hospital events (HES APC data), accident & emergency events (HES A&E data) and clinical primary care events (CPRD data). Separate variables were created for counts of outpatient events and admitted hospital events to specific consultant specialties and to possible sleep related consultants (neurology, paediatric neurology, respiratory, Ear Nose and Throat (ENT), and paediatric). Separate clinical primary care event variables were created for counts of face-to-face and telephone consultations in primary care with medical staff (qualified doctors) or any clinical staff member.

### Covariates

Covariates derived from primary care data were: sex, age, and practice size. Ethnicity was defined using primary care and linked HES data. Area-based deprivation quintiles were assigned using the Carstairs index from 2011 census mapped to practice postcode. Urban-rural categories were based on the 2011 census mapped to the patient postcode. Data management and statistical analysis were conducted using Stata MP, version 18. Programming code for data management and analysis, along with codelists and codelist checklists are publicly available[27–29][I WILL UPDATE THESE REPOSITORIES WHEN I HAVE A PREPRINT DOI]. Detailed definitions of exposures, outcomes and covariates are provided in Supplementary Appendix table 2.

## Statistical analysis

### Primary analysis

We used Poisson regression models with an offset for person-time to estimate event rate ratios (RRs) for each outcome in people in the narcolepsy and comparison group. To account for within-person correlation due to multiple observation periods and potential outcome events per individual, we used robust standard errors. Poisson models were chosen because of their stability across outcomes and time dependent analyses; the alternative of negative binomial regression (to handle overdispersion) did not consistently converge but was used for selected outcomes in sensitivity analyses (see Sensitivity Analyses, below). Models were adjusted for age and sex. To remove overinflated counts due to data linkage errors[30], we removed individuals with a crude overall event rate of >50/year from the HES APC and HES Outpatient analyses. We further discounted outcomes with less than 20 events in the narcolepsy or comparison group, using this as a pragmatic approach to avoid non-convergence and unstable estimates. Regression models were restricted to females aged 15 to 45 for hospital events with obstetricians, females aged 15 and older for events with gynaecologists, people aged 17 and under for events with paediatricians, and people aged 55 and over for events with geriatricians.

From the estimated adjusted model, we calculated predicted rates of each outcome in the narcolepsy and comparison groups by estimating average marginal (partial) effects for the observations in the data[31]. We used the same approach to estimate Rate Differences (RDs) describing the absolute difference in event rates between the narcolepsy and comparison group.

### Secondary analyses

We estimated rates, RRs, and RDs by time relative to the index date (−15 to +15 years) for all events and possible sleep-related event outcomes using interaction terms between the exposure group and year modelled with a flexible function (spline with 8 knots located at –10,-5,-2,-1,1,2,5,10 years).

We estimated stratified RRs, rates and RDs in the 5 years before and after diagnosis by sex and age group by including an interaction term for sex and categorical age in regression models for all outcomes where these models converged. Wald tests were used to test the statistical significance of interactions. We additionally estimated stratified effect estimates for all events and possible sleep related event outcomes by ethnicity, area-based deprivation quintiles, urban-rural category, calendar year and practice size quintiles.

### Sensitivity analyses

In case overdispersion affected the Poisson models, we fit negative binomial models as a sensitivity analyses for the all events outcome where these converged and produced stable dispersion estimates.

We repeated analyses for the all events outcomes using the first coded record of narcolepsy in primary care date to identify narcolepsy cases as these may be identified more accurately using this definition[26].

## RESULTS

### Study population

The narcolepsy group included 2,772 people with incident narcolepsy from a study population including 37,501,230 registration periods in GP practices (Figure 2). The matched comparison group included 13,860 individuals without narcolepsy. Two people in the comparison group were diagnosed after the index date and included in both groups. Analyses using HES Outpatient and HES A&E data included 2,753 people in the narcolepsy group and 13,764 people in the comparison group.

**Figure 2:**
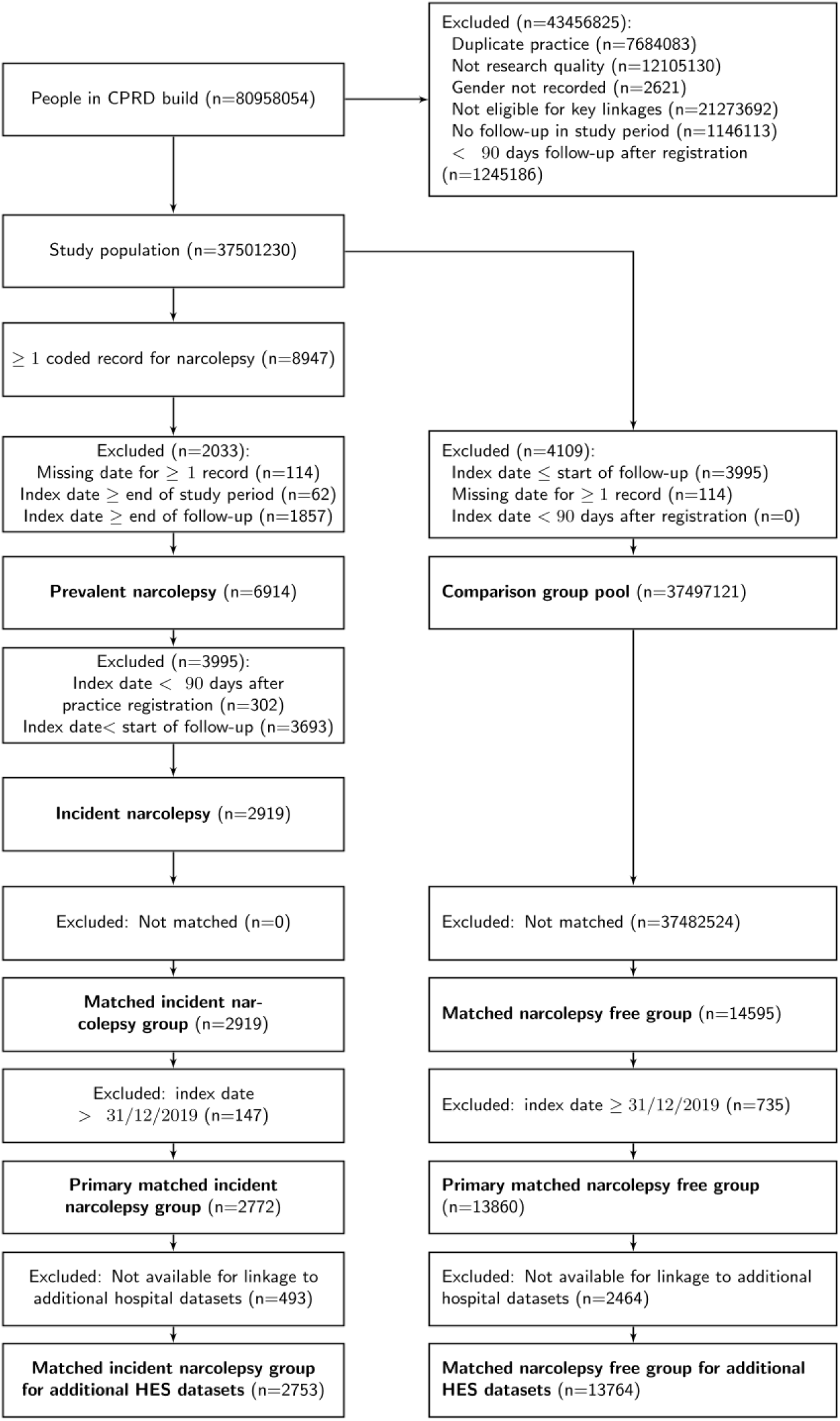
Participant Flow Chart.

### Healthcare use five years before and after diagnosis (primary analysis)

Table 1 describes the narcolepsy and comparison groups. Median (IQR) years of follow-up for the primary five years before and after analysis was 7.6 (5.6, 10.0) in the narcolepsy group and 8.0 (5.9, 10.0) in the comparison group. Median (IQR) age at index was 39.0 (25.0, 55.0) in both groups; 52.8% of individuals were female.

Table 2 presents rates, RRs and RDs for all clinical events and possible sleep related events in each setting and each primary care event type. People with narcolepsy attended each setting approximately twice as often as the comparison group in the period from five years before to five years after the index date. Per year, this amounted to an additional 1.91 (95% CI 1.76-2.05) outpatient events, 0.36 (95% CI 0.30-0.42) admitted patient care events, 0.25 (95% CI 0.22-0.29) accident and emergency events and 4.32 (95% CI 3.95-4.69) primary care events. There were 0.77 (95% CI 0.73-0.81) and 0.10 (95% CI 0.09-0.11) additional possible sleep related events in outpatient and admitted patient care settings, respectively. Most primary care consultations during the study period were face-to-face with medical staff.

**Table 2:** Rates, rate ratios and rate differences for all clinical events and possible sleep related events to each setting and each primary care visit type.

|  | Exposed group |  | Unexposed group |  | Comparison |  |
| --- | --- | --- | --- | --- | --- | --- |
| Setting / Outcome | Event count | Rate per 100 person years (95% CI) | Event count | Rate per 100 person-years (95% CI) | Rate Ratio (95% CI) | Rate Difference per 100 person-years (95% CI) |
| **Outpatient setting** |  |  |  |  |  |  |
| All events | 46824 | 3.01 (2.87, 3.15) | 90171 | 1.10 (1.06, 1.14) | 2.73 (2.58, 2.89) | 1.91 (1.76, 2.05) |
| Possible sleep-related visit | 14594 | 0.87 (0.83, 0.91) | 8352 | 0.10 (0.09, 0.10) | 9.01 (8.30, 9.79) | 0.77 (0.73, 0.81) |
| **Admitted patient care setting** |  |  |  |  |  |  |
| All events | 11795 | 0.61 (0.55, 0.67) | 25360 | 0.25 (0.22, 0.27) | 2.45 (2.14, 2.82) | 0.36 (0.30, 0.42) |
| Possible sleep-related visit | 2321 | 0.12 (0.10, 0.13) | 1531 | 0.01 (0.01, 0.02) | 7.81 (6.49, 9.40) | 0.10 (0.09, 0.11) |
| **Accident & Emergency setting** |  |  |  |  |  |  |
| All events | 7470 | 0.46 (0.43, 0.49) | 17510 | 0.21 (0.20, 0.22) | 2.21 (2.05, 2.39) | 0.25 (0.22, 0.29) |
| **Primary care setting** |  |  |  |  |  |  |
| All events | 160547 | 9.39 (9.02, 9.76) | 456074 | 5.07 (4.97, 5.18) | 1.85 (1.77, 1.93) | 4.32 (3.95, 4.69) |
| Face to face | 129624 | 9.22 (8.94, 9.50) | 389758 | 6.57 (6.47, 6.68) | 1.40 (1.36, 1.45) | 2.64 (2.36, 2.93) |
| Face to face with medical staff | 81094 | 5.63 (5.44, 5.82) | 229620 | 3.81 (3.75, 3.88) | 1.48 (1.42, 1.53) | 1.82 (1.62, 2.01) |
| Telephone/video | 21619 | 1.58 (1.44, 1.72) | 45432 | 0.78 (0.75, 0.81) | 2.03 (1.85, 2.23) | 0.80 (0.66, 0.94) |
| Telephone/video with medical staff | 19377 | 1.42 (1.28, 1.55) | 40197 | 0.69 (0.66, 0.72) | 2.06 (1.86, 2.28) | 0.73 (0.59, 0.86) |

Figure 3 and 4 plot RRs for each consultant specialty and describe RDs and rates in the narcolepsy group in the outpatient and inpatient setting, respectively. Among people with narcolepsy, the highest rates of outpatient events per year were to possible-sleep related consultants: paediatrics in people under 18 (1.56 95% CI 1.37-1.74), respiratory medicine (0.29 95% CI 0.26-0.31) and neurology (0.25 95% CI 0.23-0.27). RRs and RDs were highest for respiratory medicine (RR 17.8 95% CI 14.9-21.4; RD per year 0.27 95% CI 0.25-0.30), neurology (RR 13.0 95% CI 11.1-15.4; RD per year 0.23 95% CI 0.25-0.30) and paediatrics in children (RR 12.2 95% CI 9.84-15.0; RD per year 1.43 95% CI 1.25-1.61). There was evidence of elevated rates of events for the majority of other specialties, most notably psychiatry (RR 4.32 95% CI 2.79-6.67; RD per year 0.13 95% CI 0.08-0.18). Similar patterns were observed for admitted patient care events.

**Figure 3:**
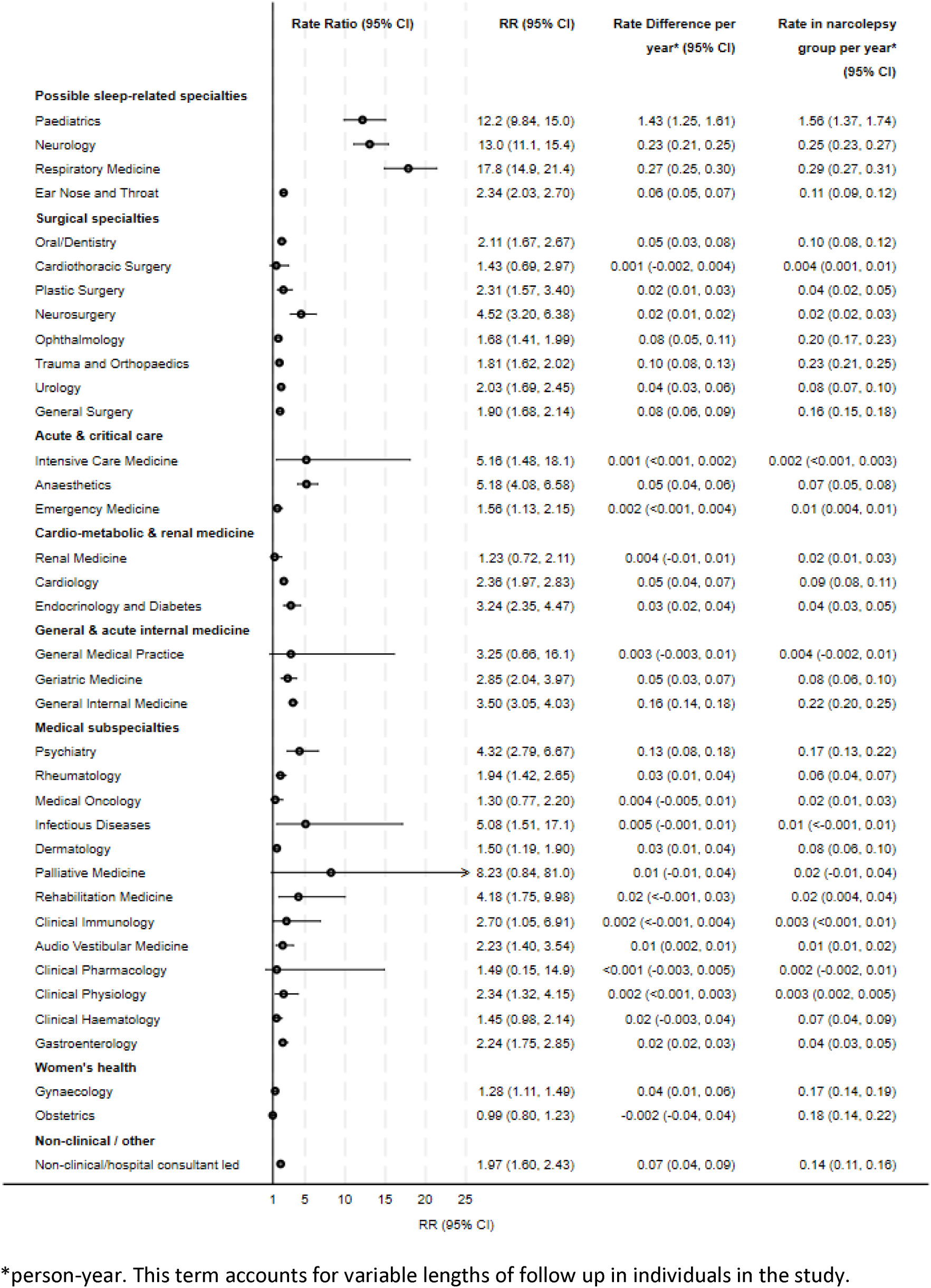
Rate Ratios and Rate Differences comparing hospital outpatient events in the narcolepsy and comparison group in the 5 years before and after index date.

**Figure 4:**
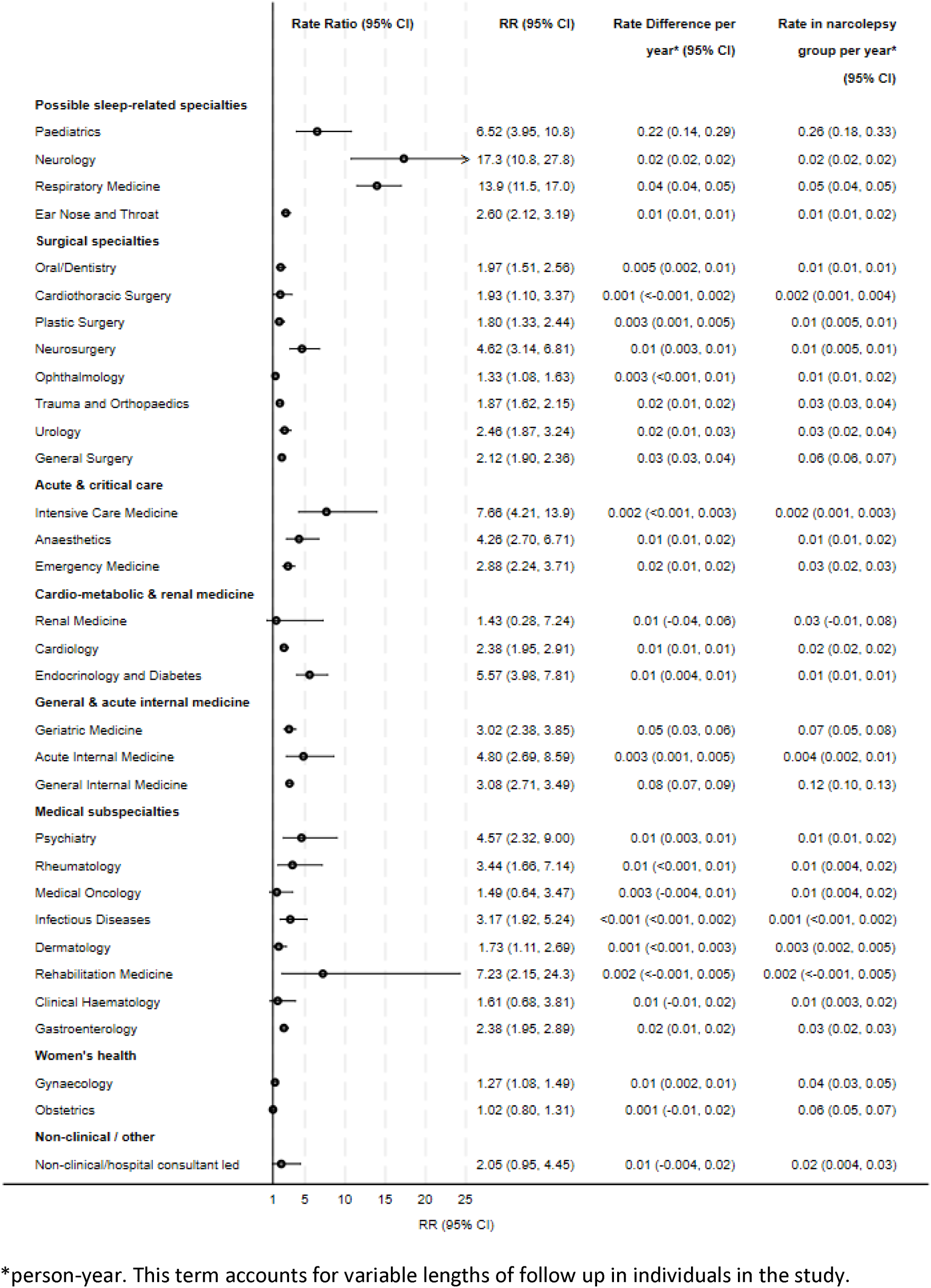
Rate Ratios and Rate Differences comparing hospital admitted patient care events in the narcolepsy and comparison group in the 5 years before and after index date.

### Healthcare use over time before and after diagnosis (secondary analysis)

Rates of each outcome in the comparison group increased modestly over time relative to the index date whereas rates in the narcolepsy group peaked at the index date (Supplementary Appendix Figure 1). RDs (Figure 5) and RRs (Supplementary Appendix Figure 2) showed increases in healthcare use in the narcolepsy group compared to the comparison group for at least 15 years before and after the index date, with peaks at the index date. RDs comparing hospital events with possible-sleep related consultants in the narcolepsy and comparison group peaked the year including and after the index date (outpatient RD per year 1.50 95% CI 1.41-1.59), decreased substantially within 5 years (outpatient RD per year 0.60 95% CI 0.53-0.67), and decreased to less than 0.5 visits per year soon after (Figure 5 and Supplementary Appendix Table 2).

**Figure 5:**
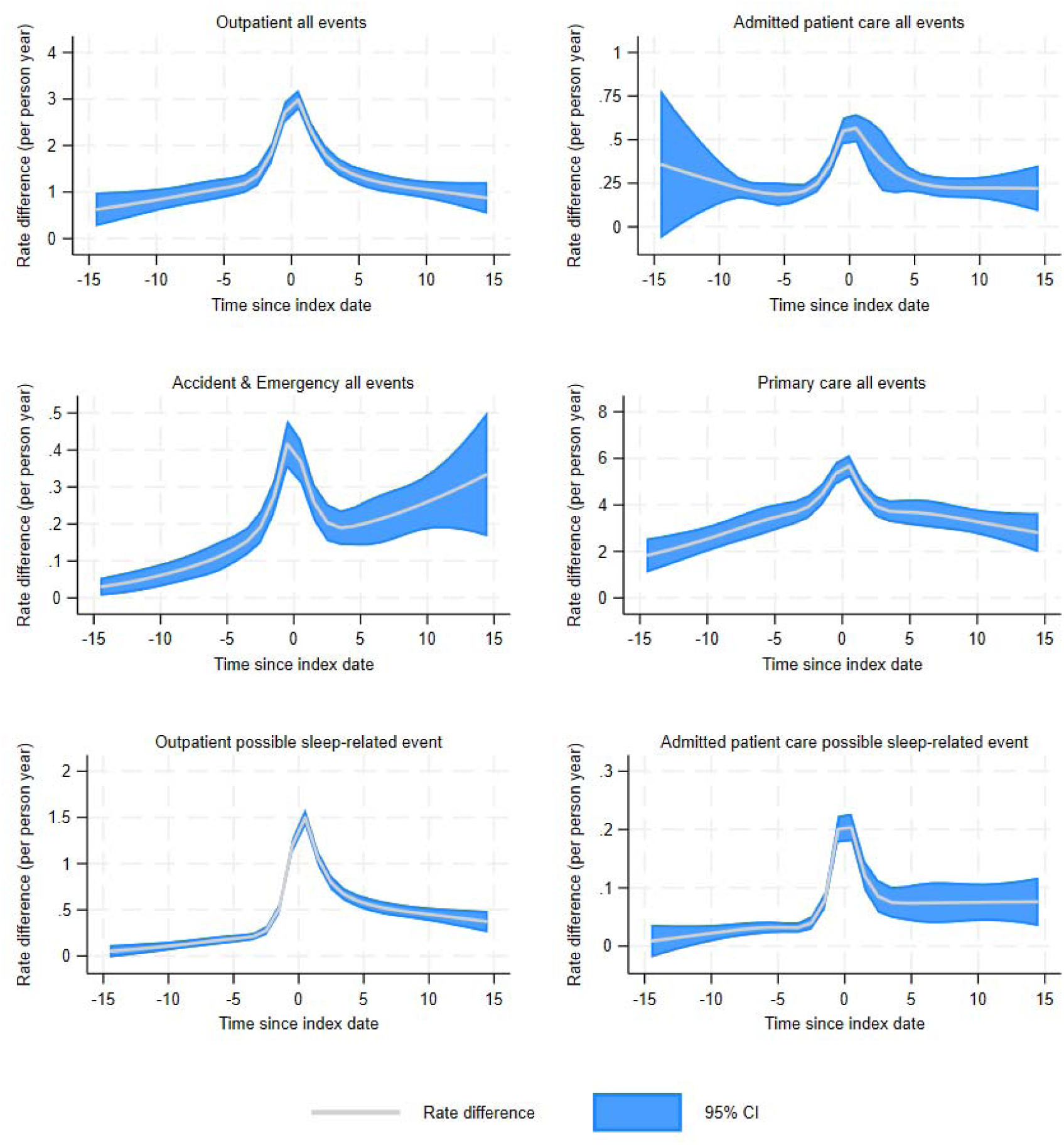
Rate Differences comparing healthcare resource use in the narcolepsy and comparison groups over time relative to the index date.

### Healthcare use stratified by demographic variables (secondary analysis)

Supplementary Appendix Tables 3a to 3f describe stratified RRs, rates and RDs for the all events and possible-sleep related event outcomes for all covariates. There is strong evidence of effect modification by age. RRs for all outpatient and admitted patient care events were highest in younger age groups and decreased with increasing age. In contrast, RDs were more similar across age groups, reflecting higher rates in the comparison group at older ages. This pattern suggests that large relative effects in younger individuals were driven by lower underlying event rates, whereas absolute differences in use were more consistent across age groups. RRs for possible-sleep related events were highest in people aged 18 to <35; RDs decreased with increasing age. There was little evidence that RRs differed by gender, ethnicity, area-based deprivation, urban/rural practices or calendar year. Where differences were observed, they were generally modest in absolute terms or stratified estimates were imprecise. Exceptions included calendar year for all outpatient events, where RRs and RDs increased over time. RRs for possible-sleep related outpatient events also varied by ethnicity, and were highest in Black people, although the corresponding RD was similar to the unstratified estimate.

There was little statistical evidence that RRs differed between males and females for most specialities (Supplementary Appendix Tables 4a and 4b). Where differences were observed (e.g. cardiology and urology), RRs were higher in females, while RDs were similar, consistent with higher comparison group event rates in males. Age differences for respiratory medicine and neurology in adults are consistent with findings for possible-sleep related event outcomes (Supplementary Appendix Tables 5a and 5b). There was evidence of differences in relative event rates for other consultant specialities where event rates in the comparison group differed between age groups.

### Sensitivity analyses

Negative binomial regression models resulted in higher RRs than Poisson models for the all event outcomes for each setting (Supplementary Appendix table 6).

RRs and RDs were similar when narcolepsy was recorded in either primary care or HES APC data (primary definition) and when narcolepsy was recorded in primary care data only (sensitivity definition) (Supplementary Appendix table 7).

## DISCUSSION

### Principal findings

This study provides estimates of healthcare use in all NHS England settings in people with narcolepsy across the patient journey including primary care, and hospital consultant specialty and over time before and after recording of narcolepsy in electronic health records. The rate of healthcare use in people with narcolepsy was approximately twice that of matched controls in the period from five years before to five years after narcolepsy was recorded, with similar relative increases observed in all NHS settings. Annually, most additional visits were to primary care following by hospital outpatient settings. The highest rates of additional hospital visits were to neurology, respiratory and paediatric specialists who are responsible for diagnosing and treating narcolepsy. However, there was strong evidence of higher rates of use for the majority of consultant specialties, most notably psychiatry. When combined, these represent a substantial proportion of additional visits. Overall healthcare use in all NHS settings and with sleep-related consultants peaked markedly when narcolepsy was recorded, followed by a substantial decline, although rates were higher for at least 15 years before and after this date.

### Strengths and weaknesses

We have used linked electronic health record and administrative datasets that are representative of the English population by age, sex, area-based deprivation and ethnicity[19,20,24,32]. These datasets are well established and supported by studies examining data quality and providing methodology guidance. This has enabled rigorous study design and transparent consideration of the strengths and limitations of this research.

Our primary definition of narcolepsy cases used coded records in primary care and HES APC data; we have previously estimated that 65.2% (95% CI 57.0% to 73.4%) of these cases were diagnosed by a specialist[33]. Using our sensitivity analysis algorithm (primary care data only), 71.0% (95% CI 58.8% to 81.3%) of narcolepsy cases were confirmed. We observed similar findings with both definitions in this and a previous study describing the incidence and prevalence of diagnosed narcolepsy in the UK[34]. Furthermore, diagnoses are commonly recorded in primary care data years after diagnosis[33]. This points to likely under-recording of narcolepsy in our data and the index date in this study should thus be interpreted with caution.

There is no identifier for sleep centres in HES data; based on our knowledge, we included neurology, respiratory, ENT and paediatric specialists in our possible-sleep related group. Measures of area-based deprivation and rural-urban status are proxies for individual socioeconomic status and rural-urban living.

We developed codelists and algorithms to identify primary care consultations in CPRD data (UPDATE AND ADD REFS WHEN I HAVE A PREPRINT DOI). These identify similar rates of primary care events in the comparison data to national data sources[35]. However, a lack of specificity of coding in the GP record may lead to over– or under-estimation of events. Our follow-up period was restricted to the period during which people were registered in the practice. Delays in delisting patients when they move out of area may lead to underestimation of primary care event rates. This would affect the comparison group more than the narcolepsy group as the latter had a coded record of narcolepsy at index.

By using Poisson models and not negative binomial regression models, we may have underestimated differences between healthcare use in people with narcolepsy compared to people without narcolepsy.

### Comparison with other studies

Our study provides a comprehensive overview of healthcare use in people with narcolepsy compared to age-sex matched controls in the general population in England. To our knowledge. this is the first full manuscript to compare primary care use and the first publication to report absolute differences in event rates, consider a wide range of hospital specialists, and to fully describe changes in healthcare use over time since diagnosis. Our findings are consistent with reported healthcare utilisation patterns among people with narcolepsy in countries as diverse as the USA [10,13,14,18], Denmark[9], Taiwan[12], as well as a smaller previous study in the UK[16]; these studies are likely to have been affected by similar data-related limitations. There was considerable variation in the methods used including case definitions, measures of effect, and time periods relative to diagnosis over which they were estimated. Two papers reported substantially greater differences in inpatient and outpatient events in people with narcolepsy compared to controls; one studied paediatric narcolepsy[11] and the second measured healthcare resource use in the year after diagnosis[15]. [14,16], [14,16][14] [9,18][9].

### Meaning of the study: possible mechanisms and implications for clinicians and policymakers

In comparison to the general population, people with narcolepsy have higher healthcare use in all NHS settings for many years before and after narcolepsy is recorded in primary care data. In England, narcolepsy is primarily diagnosed and treated in clinics led by respiratory, neurology and paediatric consultants. The decline in outpatient events to these specialists after narcolepsy is recorded suggests that people do not receive regular follow-up by specialists, matching patient reports of variable post-diagnosis support[2]. Higher rates of A&E attendances, outpatient visits and hospital admissions to multiple consultant specialists are likely to reflect the direct and indirect impacts of narcolepsy on multiple organ systems[3,7]. Psychiatric and cardio-metabolic related healthcare events, for example, may arise from narcolepsy symptoms, shared pathophysiological mechanisms, and side effects of medications[3,6,36]. A&E events may be associated with traumatic injury[37]. Undiagnosed symptoms of narcolepsy may explain some events prior to the index date[38], particularly psychiatric services[36], primary care and A&E. Earlier recognition and referral might decrease primary care and hospital events before diagnosis, while routine follow-up by sleep specialists and in primary care might prevent hospital related events caused by treatable symptoms and avoidable comorbidities. Previous research has shown that such timely diagnosis and follow-up are hindered by limited service capacity, referral guidance, and medical education about sleep disorders[39,40].

### Unanswered questions and future research

Our study shows that healthcare use is high before and after diagnosis but does not unpick the extent to which this is due to narcolepsy symptoms, associated comorbidities, side effects of medications, or where and how healthcare is delivered. Answering these questions requires individual-level data linking the timing of symptom onset, diagnosis, treatment and comorbidity to subsequent healthcare use. Population-based routinely collected data sources, including those used here, do not capture this detail. Prospective, co-ordinated, longitudinal data collection within specialist sleep services would facilitate robust epidemiological and health service research that answers these questions.

## CONCLUSION

People with narcolepsy have elevated use of healthcare across NHS settings, before diagnosis which is sustained following diagnosis. This may reflect diagnostic delay, comorbidities, and ongoing narcolepsy-related healthcare needs being met largely outside specialist care. This suggests opportunities to improve narcolepsy care through earlier diagnosis, more consistent prevention and treatment of comorbidities, and better coordination of care between specialist and non-specialist services.

## Data availability statement

This study is based in part on data from the Clinical Practice Research Datalink obtained under licence from the UK Medicines and Healthcare products Regulatory Agency. The terms of our licence to access the data prevent us from sharing individual patient data with third parties. The raw data may be requested directly from CPRD following their usual procedures.

## Ethics approval statement

The protocol for this research was approved via the Clinical Practice Research Datalink (CPRD) Research Data Governance (RDG) process (protocol 22_001887). CPRD has ethics approval from the Health Research Authority to support research using anonymised patient data. Research Ethics Committee (East Midlands—Derby, REC reference number 21/EM/0265). The study has additionally been approved by the London School of Hygiene & Tropical Medicine Ethics Committee (Ref 101296).

## Patient consent statement

CPRD supplies anonymised data for public health research; therefore, individual patient consent was not required for this study.

## Contributions

The study was conceptualised by HS (guarantor), HM, EN, SHE, MAM, KB, IES, and CWG. Methodology was developed by HS, KB and AB. HS led the investigation, formal analysis, data curation, and software development, and was responsible for funding acquisition, visualisation, and writing the original draft. All authors contributed to reviewing and editing the manuscript.

## REFERENCES

1. Ohayon MM, Dave S, Crawford S, Swick TJ, Côté ML. Prevalence of narcolepsy in representative samples of the general population of North America, Europe, and South Korea. Psychiatry Res. 2025 May 1;347:116390. doi:10.1016/j.psychres.2025.116390

2. Narcolepsy Charter | Narcolepsy UK [Internet]. 2018 [cited 2020 Jun 11]. Available from: https://www.narcolepsy.org.uk/resources/narcolepsy-charter

3. Arias-Carrión O, Ortega-Robles E, Romano P, Pineda C. Narcolepsy as an immune-associated hypothalamic encephalopathy: orexin dysfunction and implications for precision sleep medicine. Front Psychiatry. 2026 Apr 2;17. doi:10.3389/fpsyt.2026.1799520

4. Read N, Anderson K, Craig S, Kendrick A, Quinnell T, Steier J, Hare A. The Optimal Sleep Pathway: Towards better care for patients with sleep conditions [Internet]. British Sleep Society; 2025 Feb [cited 2025 Jan 4]. Available from: https://www.sleepsociety.org.uk/optimal-sleep-pathway/

5. van Someren F, Wiedemann M, Warren-Gash C, Sykorova M, Mistry H, Miller MA, Leschziner G, Nolte E, Belot A, Smith IE, Strongman H. Trends and variation in issuance of high-cost narcolepsy drugs by NHS England organisations and regions from 2019 to 2022. J Sleep Res. 2024 Aug 12;34(4):e14415. doi:10.1111/jsr.14415

6. Kwon Y, Gami AS, Javaheri S, Pressman GS, Scammell TE, Surkin LA, Zee PC. Cardiovascular Risks in People With Narcolepsy: Expert Panel Consensus Recommendations. J Am Heart Assoc. 2024 Aug 20;13(16):e035168. doi:10.1161/JAHA.124.035168

7. Gudka S, Haynes E, Scotney J, Mukherjee S, Frenkel S, Sivam S, Swieca J, Chamula K, Cunnington D, Saini B. Narcolepsy: Comorbidities, complexities and future directions. Sleep Med Rev. 2022 Oct 1;65:101669. doi:10.1016/j.smrv.2022.101669

8. NHS Englandlll» Shared Care Protocols [Internet]. [cited 2026 Jul 21]. Available from: https://www.england.nhs.uk/medicines-2/regional-medicines-optimisation-committees-advice/shared-care-protocols/

9. Jennum P, Ibsen R, Petersen ER, Knudsen S, Kjellberg J. Health, social, and economic consequences of narcolepsy: A controlled national study evaluating the societal effect on patients and their partners. Sleep Med. 2012 Sep;13(8):1086–93. doi:10.1016/j.sleep.2012.06.006

10. Black J, Reaven NL, Funk SE, McGaughey K, Ohayon M, Guilleminault C, Ruoff C, Mignot E. The Burden of Narcolepsy Disease (BOND) study: Health-care utilization and cost findings. Sleep Med. 2014;15(5):522–9. doi:10.1016/j.sleep.2014.02.001 PubMed PMID: 24768358.

11. Carls G, Reddy SR, Broder MS, Tieu R, Villa KF, Profant J, Halbower AC. Burden of disease in pediatric narcolepsy: a claims-based analysis of health care utilization, costs, and comorbidities. Sleep Med. 2020 Feb 1;66:110–8. doi:10.1016/j.sleep.2019.08.008

12. Huang YS, Chin WC, Chung IH, Roan TY, Chang CJ, Juang HT, Chang SC, Ghosh S, Crawford S, Lin HL. The prevalence, incidence, and impact of narcolepsy and idiopathic hypersomnia in Taiwan: comparison between the National Health Insurance Research Claims Database and a hospital cohort database. Sleep. 2025 Nov 1;48(11):zsaf132. doi:10.1093/sleep/zsaf132

13. Saad R, Markt SC, Lillaney P, Profant DA, Fuller DS, Poole EM, Alvord T, Prince P, Desai S, Whalen M, Ni W, Black J. The Clinical and Economic Burden of Idiopathic Hypersomnia and Narcolepsy: A United States Claims-Based Analysis. Nat Sci Sleep. 2025;17:1809–23. doi:10.2147/NSS.S498213 PubMed PMID: 40800589; PubMed Central PMCID: PMC12341555.

14. Flores NM, Villa KF, Black J, Chervin RD, Witt EA. The Humanistic and Economic Burden of Narcolepsy. J Clin Sleep Med. 2016 Mar 15;12(03):401–7. doi:10.5664/jcsm.5594

15. Kadotani H, Matsuo M, Tran L, Parsons VL, Maguire A, Ghosh S, Crawford S, Dave S. Healthcare burden of narcolepsy in Japan: A retrospective analysis of health insurance claims from the Japan Medical Data Center. Sleep Med. 2025 Mar;127:64–72. doi:10.1016/j.sleep.2025.01.003 PubMed PMID: 39809067.

16. Crawford S, Azpeitia Á, Ghosh S, O’Reilly J, Podmore B, Qizilbash N. Healthcare burden of narcolepsy in the United Kingdom: a cohort study from the CPRD and HES databases. Sleep Med. 2024 Feb 1;Abstracts from the 17th World Sleep Congress115:S216. doi:10.1016/j.sleep.2023.11.601

17. Hobbs FDR, Bankhead C, Mukhtar T, Stevens S, Perera-Salazar R, Holt T, Salisbury C. Clinical workload in UK primary care: a retrospective analysis of 100 million consultations in England, 2007–14. The Lancet. 2016 Jun 4;387(10035):2323–30. doi:10.1016/S0140-6736(16)00620-6 PubMed PMID: 27059888.

18. Villa KF, Reaven NL, Funk SE, McGaughey K, Black J. Changes in Medical Services and Drug Utilization and Associated Costs After Narcolepsy Diagnosis in the United States. Am Health Drug Benefits. 2018;11(3):137–45. PubMed PMID: 29910845.

19. Wolf A, Dedman D, Campbell J, Booth H, Lunn D, Chapman J, Myles P. Data resource profile: Clinical Practice Research Datalink (CPRD) Aurum. Int J Epidemiol. 2019 Mar 11;48(6):1740–1740g. doi:10.1093/ije/dyz034

20. Sanchez-Santos MT, Axson EL, Dedman D, Delmestri A. Data Resource Profile Update: CPRD GOLD. Int J Epidemiol. 2025 Aug 1;54(4):dyaf077. doi:10.1093/ije/dyaf077

21. Padmanabhan S, Carty L, Cameron E, Ghosh RE, Williams R, Strongman H. Approach to record linkage of primary care data from Clinical Practice Research Datalink to other health-related patient data: overview and implications. Eur J Epidemiol. 2019 Jan 15;34(1):91–9. doi:10.1007/s10654-018-0442-4 PubMed PMID: 30219957.

22. Herbert A, Wijlaars L, Zylbersztejn A, Cromwell D, Hardelid P. Data Resource Profile: Hospital Episode Statistics Admitted Patient Care (HES APC). Int J Epidemiol. 2017 Aug 1;46(4):1093–1093i. doi:10.1093/ije/dyx015 PubMed PMID: 28338941.

23. Gallagher AM, Dedman D, Padmanabhan S, Leufkens HGM, de Vries F. The accuracy of date of death recording in the Clinical Practice Research Datalink GOLD database in England compared with the Office for National Statistics death registrations. Pharmacoepidemiol Drug Saf. 2019 May 1;28(5):563–9. doi:10.1002/pds.4747 PubMed PMID: 30908785.

24. Mahadevan P, Harley M, Fordyce S, Hodgson S, Ghosh R, Myles P, Booth H, Axson E. Completeness and representativeness of small area socioeconomic data linked with the UK Clinical Practice Research Datalink (CPRD). J Epidemiol Community Health. 2022 Oct 1;76(10):880–6. doi:10.1136/jech-2022-219200 PubMed PMID: 35902219.

25. Mansfield KE, Mathur R, Tazare J, Henderson AD, Mulick AR, Carreira H, Matthews AA, Bidulka P, Gayle A, Forbes H, Cook S, Wong AYS, Strongman H, Wing K, Warren-Gash C, Cadogan SL, Smeeth L, Hayes JF, Quint JK, McKee M, Langan SM. Indirect acute effects of the COVID-19 pandemic on physical and mental health in the UK: a population-based study. Lancet Digit Health. 2021 Apr 1;3(4):e217–30. doi:10.1016/S2589-7500(21)00017-0 PubMed PMID: 33612430.

26. Strongman H, Eriksson SH, Asare K, Miller MA, Sykorova M, Mistry H, Veighey K, Warren-Gash C, Bhaskaran K. Validation of algorithms identifying diagnosed Obstructive Sleep Apnoea and narcolepsy in coded primary care and linked hospital activity data in England [Internet]. medRxiv; 2025 [cited 2025 Mar 25]. p. 2025.03.10.25323660. Available from: https://www.medrxiv.org/content/10.1101/2025.03.10.25323660v1 doi:10.1101/2025.03.10.25323660

27. Strongman H. Chronology of healthcare resource use and comorbidities in people with obstructive sleep apnoea and narcolepsy before and after diagnosis: a descriptive study: Github repository v1.01 Descriptive epidemiology [Internet]. Zenodo; 2025 [cited 2025 Jul 31]. Available from: https://zenodo.org/records/15756446 doi:10.5281/zenodo.15756446

28. Strongman H, Eriksson SH, Quinnell T, Mathur R, Lyons A, Wong A. Codelists for “Chronology of healthcare resource use and comorbidities in people with obstructive sleep apnoea and narcolepsy before and after diagnosis: a descriptive study” [Internet]. London, United Kingdom: London School of Hygiene & Tropical Medicine; 2025 [cited 2025 Jul 17]. Available from: https://datacompass.lshtm.ac.uk/id/eprint/4742/ doi:10.17037/DATA.00004742

29. Matthewman J, Andresen K, Suffel A, Lin LY, Schultze A, Tazare J, Bhaskaran K, Williamson E, Costello R, Quint J, Strongman H. Checklist and guidance on creating codelists for routinely collected health data research [version 2; peer review: 3 approved]. NIHR Open Res. 2024 Sep 18;4:20. doi:10.3310/nihropenres.13550.2

30. Doidge JC, Harron KL. Reflections on modern methods: linkage error bias. Int J Epidemiol. 2019 Dec 1;48(6):2050–60. doi:10.1093/ije/dyz203

31. StataCorp. margins: Marginal means, predictive margins, and marginal effects [Internet]. College Station, TX: StataCorp LLC; 2025. Available from: https://www.stata.com/manuals/rmargins.pdf

32. Shiekh SI, Harley M, Ghosh RE, Ashworth M, Myles P, Booth HP, Axson EL. Completeness, agreement, and representativeness of ethnicity recording in the United Kingdom’s Clinical Practice Research Datalink (CPRD) and linked Hospital Episode Statistics (HES). Popul Health Metr. 2023 Mar 14;21(1):3. doi:10.1186/s12963-023-00302-0 PubMed PMID: 36918866; PubMed Central PMCID: PMC10013294.

33. Strongman H, Eriksson SH, Asare K, Miller MA, Sýkorová M, Mistry H, Veighey K, Warren-Gash C, Bhaskaran K. Validation of algorithms identifying diagnosed obstructive sleep apnoea and narcolepsy in coded primary care and linked hospital activity data in England. Sleep Epidemiol. 2025 Dec 1;5:100110. doi:10.1016/j.sleepe.2025.100110

34. Strongman H, Sykorova M, Yu YTN, Belot A, Mistry H, Nolte E, Eriksson SH, Miller MA, Bhaskaran K, Smith IE, Warren-Gash C. Incidence and prevalence of obstructive sleep apnoea and narcolepsy in the UK: a population-based descriptive study. Thorax. 2026 Feb 5;thorax-2025-223863. doi:10.1136/thorax-2025-223863 PubMed PMID: 41644140.

35. NHS England Digital [Internet]. [cited 2026 Jun 22]. Appointments in General Practice. Available from: https://digital.nhs.uk/data-and-information/publications/statistical/appointments-in-general-practice

36. Morse AM, Sanjeev K. Narcolepsy and Psychiatric Disorders: Comorbidities or Shared Pathophysiology? Med Sci. 2018 Feb 15;6(1):16. doi:10.3390/medsci6010016 PubMed PMID: 29462876; PubMed Central PMCID: PMC5872173.

37. Zheng Y, Fukasawa T, Masuda S, Takeuchi M, Kawakami K. Narcolepsy and risk of traumatic injury: a population-based matched cohort study. J Clin Sleep Med JCSM Off Publ Am Acad Sleep Med. 2024 Oct 1;20(10):1657–62. doi:10.5664/jcsm.11236 PubMed PMID: 38913343; PubMed Central PMCID: PMC11446133.

38. Thorpy MJ, Krieger AC. Delayed diagnosis of narcolepsy: characterization and impact. Sleep Med. 2014 May 1;15(5):502–7. doi:10.1016/j.sleep.2014.01.015

39. Romiszewski S, May FEK, Homan EJ, Norris B, Miller MA, Zeman A. Medical student education in sleep and its disorders is still meagre 20 years on: A cross-sectional survey of UK undergraduate medical education. J Sleep Res. 2020 Dec 12;29(6):e12980. doi:10.1111/jsr.12980

40. Sykorova M, Someren F van, Veighey K, Nolte E, Warren-Gash C, Miller MA, Eriksson SH, Smith IE, Strongman H. Factors influencing English general practitioners referrals to specialist sleep services: a qualitative study using the COM-B model [Internet]. medRxiv; 2025 [cited 2025 Oct 29]. p. 2025.10.26.25338808. Available from: https://www.medrxiv.org/content/10.1101/2025.10.26.25338808v1 doi:10.1101/2025.10.26.25338808

