## Supplementary Appendix for "Patterns of healthcare use in people with narcolepsy: a population-based cohort study in England"

**SA Table 1**: **Data sources, versions and DOIs**

| **Database** | **Version** | **DOI** |
| --- | --- | --- |
| CPRD Aurum | 2022.03.001 | <https://doi.org/10.48329/MY9S-4X08> |
| CPRD GOLD | 2022.03.001 | <https://doi.org/10.48329/5RHX-ER29> |
| CPRD GOLD HES APC | 2022.01.001 | <https://doi.org/10.48329/fagm-ez75> |
| CPRD Aurum HES APC | 2022.01.001 | <https://doi.org/10.48329/vagx-9d96> |
| CPRD GOLD HES Outpatient | 2021.08.001 | <https://doi.org/10.48329/cp5e-7790> |
| CPRD Aurum HES Outpatient | 2021.08.001 | <https://doi.org/10.48329/7hm3-gt75> |
| CPRD GOLD HES A&E | 2021.08.001 | <https://doi.org/10.48329/xtwg-gp32> |
| CPRD Aurum HES A&E | 2021.08.001 | <https://doi.org/10.48329/2QY5-XX60> |
| CPRD GOLD ONS deaths | 2022.01.001 | <https://doi.org/10.48329/wq7v-x832> |
| CPRD Aurum ONS deaths | 2022.01.001 | <https://doi.org/10.48329/Q34F-F505> |
| CPRD GOLD Small Area data (practice) – Carstairs Index | 2022.01.001 | <https://doi.org/10.48329/aep7-aq95> |
| CPRD Aurum Small Area data (practice) – Carstairs Index | 2022.01.001 | <https://doi.org/10.48329/3saw-rz63> |
| CPRD GOLD Small Area data (patient) – rural-urban classification | 2022.01.001 | <https://doi.org/10.48329/y101-0w14> |
| CPRD Aurum Small Area data (patient) – rural-urban classification | 2022.01.001 | <https://doi.org/10.48329/aytt-h222> |

**SA Table 2: Variable definitions**

| **Variable** | **Type** | **Categories/units** | **Data source** | **Definition** |
| --- | --- | --- | --- | --- |
| **Exposure (see Figure 1 in manuscript)** | | | | |
| **Outcomes** | | | | |
| Outpatient event (all events) | Count | Number of specialties visited in one day | HES Outpatient | Outpatient appointments with a recorded consultant specialty (mainspef in source data) and attended by patient (attendsumm = 1). Outpatient visits to the same specialty on the same day were counted as one visit. |
| Outpatient event date | integer | Day | HES Outpatient | Appointment date (apptdate in source data) |
| Outpatient event to consultant specialty | Multiple integer variables | 0/1 | HES Outpatient | Outpatient visit including at least one episode led by specific consultant specialty. Consultant specialties were identified using a modified version of mainspef variable from source file (see footnotes). |
| Outpatient event to sleep-related specialty | binary | 0/1 | HES Outpatient | Outpatient visit including at least one episode led by consultant who may diagnose and treat sleep disorders (mainspef = 400 Neurology, 401 Paediatric neurology 421 neurophysiology, 340 respiratory, 120 ENT, 420 paediatric |
| Admitted patient care hospitalisation | Binary | 0/1 | HES APC | Admitted patient care hospitalisation (hospitalisations/spells in source data) with recorded consultant specialty (mainspef in source data). Admissions on the same or subsequent days were combined and counted as one visit. |
| Admitted patient care event date | integer | Day | HES APC | Admission date (admidate in source data) |
| Admitted patient care event to specialty | Multiple integer variables | 0/1 | HES APC | Admitted patient care hospitalisation including at least one episode led by specific consultant specialty. Consultant specialties were identified using a modified version of mainspef variable from source file (see footnotes). |
| Hospital event with sleep related consultant | binary | 0/1 | HES APC | Admitted patient care hospitalisation including at least one episode led by consultant who may diagnose and treat sleep disorders (mainspef = 400 Neurology, 401 Paediatric neurology 421 neurophysiology, 340 respiratory, 120 ENT, 420 paediatric |
| Accident & Emergency event | binary | 0/1 | HES Accident & Emergency | Accident and Emergency visit. Visits on same day counted once. |
| Accident & Emergency event date | integer | Day | HES Accident & Emergency | Accident & emergency visit date (arrivaldate in raw data). |
| Emergency department event | binary | 0/1 | HES Accident & Emergency | Visit to emergency department (aedepttype = 1 in source data). Visits on same day counted once. |
| Primary care event | count | Number of primary care consultations in one day | CPRD | Defined using consultation type and staff group codelists.  This includes face to face, telephone/video, online messaging and unspecified patient facing consultations (consmode >0 and <=5) with qualified doctors, nursing staff, allied healthcare professionals and other health care roles (staff group >1). Records with missing consultation type or staff group and consultations with the same staff group /consmode/event date were excluded. Events with different consultation mode/staff group on same day counted as unique events.  Where consmode differed between the consourceid and consmedcode in CPRD Aurum the most specific code was retained unless patient facing consultation types differed in which case consmode was set to patient facing NOS (consmode = 5). |
| Face to face primary care event | count | Number of face to face primary care consultations in one day | CPRD | Face to face visit (consmode = 1) with qualified doctors, nursing staff, allied healthcare professionals and other health care roles (staff group >1). Events with different staff groups counted as unique events. |
| Face to face event with medical staff in primary care | binary | 0/1 | CPRD | Face to face visit (consmode = 1) with qualified doctors (staff group = 2) in primary care. Duplicate visits on same day counted once. |
| Telephone event in primary care | count | Number of telephone consultations in one day | CPRD | Telephone consultation (consmode = 3) with qualified doctors, nursing staff, allied healthcare professionals and other health care roles (staff group >1). Events with different staff groups counted as unique events. |
| Telephone event in primary care with medical stafff | binary | 0/1 | CPRD | Telephone consultation (consmode = 3) with qualified doctors (staff group = 2) in primary care. Duplicate visits on same day counted once. |
| **Covariates** |  |  |  |  |
| Sex | binary | male, female | CPRD | sex recorded in GP practice |
| Age at index (for matching) | continuous | years | CPRD | age at index. Formular = 01/07/birthyear - indexdate |
| Age (for adjustment) | continuous | Years | CPRD | Age at midpoint of episode estimated with date of birth imputed as 01/07/birthyear. 3-knotted cubic spline used for adjustment. |
| Age (for stratification) | categorical | < 18, 10-year categories, >85 | CPRD | see above |
| Year relative to index | Categorical | Integers between – 15 and +15 years | N/A | The data is timesplit by year relative to diagnosis. -1 = year prior to diagnosis, +1 = year after diagnosis. There is no year 0. |
| Year relative to index splines | continuous | 8-knotted restricted cubic spline (-10, -5,-2,-1, 1, 2, 5,10 years) | N/A |  |
| Calendar year at index and in years relative to index | continuous | Integer values for each year |  |  |
| Categorical calendar year at index and in years relative to index | categorical | <2000, 2000-2004, 2005-2009, 2010-2014, 2015-2019 | N/A | Year at sleep disorder diagnosis / Year of follow-up row |
| Ethnicity | categorical | White, South Asian, Black, Other, Mixed | CPRD/ HES (all) | Derived using the most commonly recorded ethnicity in primary care data (or latest if equally common). Unknown and missing values replaced with most commonly reported ethnicity in HES where available. See Github site for more detail. |
| Practice area-based deprivation quintiles | categorical | 1 (least deprived), 2, 3, 4, 5 (most deprived) | Carstairs Index linked to the 2011 census | linked to practice postcode. Quintiles are estimated by CPRD using national Carstairs data. |
| Patient urban-rural status | binary | urban, rural | Rural Urban Classifications (RUC) for England and Wales for 2011 census population | linked to patient postcode. Missing if practice not linked to rural/urban data in census. |
| Practice size | continuous | registered patients | CPRD | Practice size for each practice estimated using full study population for incidence/prevalence study in mid-2019 or the year prior to the index date for practices leaving CPRD prior to mid-2019. |

The following mainspef consultant codes were grouped:

**Cardiology:** 320 Cardiology 321 Paediatric Cardiology

**General surgery:** 100 General Surgery 171 Paediatric Surgery

**Neurology:** 400 Neurology 401 Clinical Neurophysiology 421 Paediatric Neurology (401 is very rare in the unexposed group)

**Oral/dentistry**: 140 Oral Surgery ; 141 Restorative Dentistry 142 Paediatric Dentistry 143 Orthodontics 145 Oral and Maxillofacial Surgery 146 Endodontics 147 Periodontics 148 Prosthodontics 149 Surgical Dentistry 450 Dental Medicine 451 Special Care Dentistry 601 General Dental Practice 902 Community Health Services Dental 904 Public Health Dental

**Psychiatry**: 700 Learning Disability 710 Adult Mental Illness 711 Child and Adolescent Psychiatry 712 Forensic Psychiatry 713 Medical Psychotherapy 715 Old Age Psychiatry

**Non-clinical/consultant led**: 199 Non-UK Provider - Treatment Mainly Surgical 312 Clinical Cytogenetics and Molecular Genetics (Retired 1 April 2010) 499 Non-UK Provider - Treatment Mainly Medical 504 Community Sexual and Reproductive Health 510 Antenatal Clinic (Retired 1 April 2004) 520 Postnatal Clinic (Retired 1 April 2004)560 Midwifery 610 Maternity Function (Retired 1 April 2004) 620 Other than maternity

800 Clinical Oncology 810 Radiology 820 General Pathology 821 Blood Transfusion 822 Chemical Pathology 823 Haematology 824 Histopathology 830 Immunopathology 831 Medical Microbiology and Virology 832 Neuropathology (Retired 1 April 2004) 833 Medical Microbiology 834 Medical Virology 900 Community Medicine 901 Occupational Medicine 903 Public Health Medicine 950 Nursing 960 Allied Health Professional 990 Joint Consultant Clinics (Retired 1 April 2004).

See <https://digital.nhs.uk/data-and-information/publications/statistical/hospital-outpatient-activity/outpatient-data-quality-report> for more information above the above codes. Recording may vary between trusts and over time.

**SA Table 2 Rates, Rate Ratios and Rate Differences at specific time points relative to the index date**

|  | **Narcolepsy group** | | **Comparison group** | | **Comparison** | |
| --- | --- | --- | --- | --- | --- | --- |
| Setting/Outcome / Event type / Year relative to index date | Event count | Rate per person-year  (95% CI) | Event count | Rate per person-year  (95% CI) | Relative Rate  (95% CI) | Rate Difference per person-year  (95% CI) |
| **Outpatient** | | | | | | |
| **All events** | | | | | | |
| year prior | 7266 | 3.84 (3.62, 4.07) | 10724 | 1.13 (1.08, 1.19) | 3.40 (3.15, 3.66) | 2.71 (2.49, 2.94) |
| year after (includes index) | 8221 | 4.13 (3.92, 4.33) | 11624 | 1.14 (1.09, 1.20) | 3.61 (3.37, 3.87) | 2.98 (2.77, 3.20) |
| 4 to 5 years after | 3473 | 2.56 (2.39, 2.74) | 8960 | 1.17 (1.12, 1.23) | 2.18 (2.01, 2.37) | 1.39 (1.21, 1.57) |
| **Possible sleep-related events** | | | | | | |
| year prior | 2752 | 1.29 (1.21, 1.37) | 1055 | 0.10 (0.09, 0.11) | 12.9 (11.5, 14.6) | 1.19 (1.11, 1.27) |
| year after (includes index) | 3565 | 1.60 (1.51, 1.69) | 1058 | 0.10 (0.08, 0.11) | 16.8 (14.9, 18.9) | 1.50 (1.41, 1.59) |
| 4 to 5 years after | 914 | 0.70 (0.63, 0.77) | 719 | 0.10 (0.09, 0.11) | 6.89 (6.08, 7.82) | 0.60 (0.53, 0.67) |
| **Admitted Patient Care** | | | | | | |
| **All events** | | | | | | |
| year prior | 1931 | 0.79 (0.71, 0.86) | 2911 | 0.24 (0.22, 0.25) | 3.33 (2.94, 3.76) | 0.55 (0.47, 0.63) |
| year after (includes index) | 2063 | 0.82 (0.74, 0.89) | 3226 | 0.25 (0.23, 0.28) | 3.24 (2.82, 3.74) | 0.57 (0.49, 0.65) |
| 4 to 5 years after | 751 | 0.52 (0.45, 0.59) | 2013 | 0.24 (0.23, 0.26) | 2.13 (1.82, 2.48) | 0.27 (0.20, 0.35) |
| **Possible sleep-related events** | | | | | | |
| year prior | 557 | 0.22 (0.19, 0.24) | 210 | 0.02 (0.01, 0.02) | 13.8 (10.6, 17.9) | 0.20 (0.18, 0.22) |
| year after (includes index) | 545 | 0.22 (0.20, 0.24) | 197 | 0.02 (0.01, 0.02) | 13.9 (11.3, 17.1) | 0.20 (0.18, 0.23) |
| 4 to 5 years after | 142 | 0.09 (0.06, 0.12) | 109 | 0.02 (0.01, 0.02) | 5.36 (3.83, 7.50) | 0.07 (0.04, 0.10) |
| **Accident & Emergency (all events)** | | | | | | |
| year prior | 1310 | 0.64 (0.58, 0.70) | 2286 | 0.23 (0.21, 0.24) | 2.85 (2.53, 3.21) | 0.42 (0.35, 0.48) |
| year after (includes index) | 1246 | 0.60 (0.54, 0.65) | 2394 | 0.23 (0.21, 0.24) | 2.63 (2.35, 2.95) | 0.37 (0.31, 0.43) |
| 4 to 5 years after | 538 | 0.42 (0.37, 0.48) | 1574 | 0.23 (0.22, 0.24) | 1.84 (1.62, 2.10) | 0.19 (0.14, 0.25) |
| **Primary care consultations (all events)** | | | | | | |
| year prior | 22655 | 10.5 (10.1, 11.0) | 56385 | 5.19 (5.06, 5.33) | 2.03 (1.93, 2.13) | 5.34 (4.86, 5.83) |
| year after (includes index) | 23278 | 10.8 (10.3, 11.3) | 57101 | 5.13 (4.99, 5.26) | 2.11 (2.01, 2.21) | 5.67 (5.20, 6.15) |
| 4 to 5 years after | 11365 | 9.10 (8.59, 9.60) | 39132 | 5.40 (5.27, 5.52) | 1.69 (1.59, 1.79) | 3.70 (3.18, 4.22) |

**SA Tables 3a to 3f: Rate Ratios, Rates and Rate Differences for all events and possible-sleep related events stratified by demographic covariates**

**SA3a Hospital outpatient events (all events)**

| **Covariate / level** | **Rate Ratio (95% CI)** | **Rate per person-year in narcolepsy group** | **Rate per person-year in comparison group** | **Rate Difference per person-year (95% CI)** | **Interaction p-value (95% CI)** |
| --- | --- | --- | --- | --- | --- |
| **Age group** |  |  |  |  |  |
| <18 | 4.60 (3.93, 5.40) | 2.70 (2.40, 2.99) | 0.59 (0.52, 0.65) | 2.11 (1.81, 2.42) | <0.001 |
| 18 to <35 | 3.05 (2.76, 3.35) | 2.03 (1.88, 2.19) | 0.67 (0.63, 0.71) | 1.37 (1.21, 1.53) |  |
| 35 to <55 | 3.10 (2.78, 3.46) | 2.91 (2.68, 3.14) | 0.94 (0.87, 1.01) | 1.97 (1.73, 2.22) |  |
| 55 to <75 | 2.30 (2.06, 2.58) | 3.55 (3.21, 3.88) | 1.54 (1.45, 1.63) | 2.01 (1.66, 2.36) |  |
| 75 plus | 1.64 (1.40, 1.93) | 3.86 (3.30, 4.43) | 2.35 (2.20, 2.51) | 1.51 (0.92, 2.10) |  |
| **Gender** |  |  |  |  |  |
| Male | 2.81 (2.56, 3.08) | 2.66 (2.47, 2.85) | 0.95 (0.89, 1.00) | 1.71 (1.51, 1.91) | 0.43 |
| Female | 2.68 (2.48, 2.88) | 3.30 (3.09, 3.50) | 1.23 (1.18, 1.28) | 2.07 (1.85, 2.28) |  |
| **Ethnicity** |  |  |  |  |  |
| White | 2.58 (2.42, 2.74) | 2.99 (2.84, 3.14) | 1.16 (1.12, 1.20) | 1.83 (1.67, 1.99) | 0.83 |
| South Asian | 2.42 (1.79, 3.29) | 3.49 (2.59, 4.40) | 1.44 (1.21, 1.68) | 2.05 (1.12, 2.99) |  |
| Black | 2.87 (2.24, 3.67) | 3.70 (3.11, 4.28) | 1.29 (1.04, 1.54) | 2.41 (1.78, 3.04) |  |
| Other | 2.12 (1.29, 3.48) | 2.62 (1.64, 3.60) | 1.23 (0.83, 1.64) | 1.38 (0.33, 2.44) |  |
| Mixed | 2.50 (1.78, 3.51) | 3.12 (2.38, 3.87) | 1.25 (0.94, 1.56) | 1.87 (1.07, 2.68) |  |
| **Area-based deprivation** |  |  |  |  |  |
| 1 (least deprived) | 2.74 (2.30, 3.27) | 2.84 (2.41, 3.28) | 1.04 (0.94, 1.13) | 1.81 (1.36, 2.25) | 0.59 |
| 2 | 2.63 (2.30, 3.01) | 2.75 (2.44, 3.05) | 1.04 (0.97, 1.12) | 1.70 (1.39, 2.02) |  |
| 3 | 2.58 (2.33, 2.87) | 2.66 (2.45, 2.88) | 1.03 (0.96, 1.10) | 1.63 (1.41, 1.86) |  |
| 4 | 2.96 (2.60, 3.36) | 3.34 (2.97, 3.70) | 1.13 (1.05, 1.20) | 2.21 (1.84, 2.58) |  |
| 5 (most deprived) | 2.73 (2.42, 3.08) | 3.35 (3.04, 3.65) | 1.23 (1.13, 1.32) | 2.12 (1.80, 2.44) |  |
| **Urban-Rural practice** |  |  |  |  |  |
| urban | 2.71 (2.55, 2.89) | 3.08 (2.93, 3.24) | 1.14 (1.09, 1.18) | 1.95 (1.79, 2.11) | 0.60 |
| rural | 2.83 (2.44, 3.30) | 2.53 (2.20, 2.86) | 0.89 (0.82, 0.96) | 1.63 (1.30, 1.97) |  |
| **Calendar year*** |  |  |  |  |  |
| 1998-1999 | 1.16 (0.75, 1.79) | 1.03 (0.67, 1.39) | 0.89 (0.66, 1.12) | 0.14 (-0.28, 0.57) | <0.001 |
| 2000-2004 | 2.21 (1.93, 2.53) | 2.03 (1.80, 2.25) | 0.92 (0.85, 0.99) | 1.11 (0.87, 1.35) |  |
| 2005-2009 | 2.66 (2.39, 2.97) | 2.66 (2.43, 2.89) | 1.00 (0.93, 1.07) | 1.66 (1.42, 1.90) |  |
| 2010-2014 | 2.65 (2.39, 2.95) | 3.00 (2.75, 3.25) | 1.13 (1.06, 1.20) | 1.87 (1.61, 2.13) |  |
| 2015-2019 | 3.08 (2.76, 3.43) | 3.85 (3.51, 4.19) | 1.25 (1.17, 1.33) | 2.60 (2.25, 2.95) |  |

*includes 1998-1999 because index date is within coverage period but follow-up is sometimes earlier.

**SA3b Hospital outpatient events (Possible sleep related)**

| **Covariate / level** | **Rate Ratio (95% CI)** | **Rate per person-year in narcolepsy group** | **Rate per person-year in comparison group** | **Rate Difference per person-year (95% CI)** | **Interaction p-value (95% CI)** |
| --- | --- | --- | --- | --- | --- |
| **Age group** |  |  |  |  |  |
| <18 | 10.4 (8.65, 12.4) | 1.87 (1.68, 2.07) | 0.18 (0.15, 0.21) | 1.69 (1.50, 1.89) | <0.001 |
| 18 to <35 | 16.8 (14.3, 19.7) | 0.78 (0.72, 0.84) | 0.05 (0.04, 0.05) | 0.73 (0.67, 0.79) |  |
| 35 to <55 | 9.95 (8.61, 11.5) | 0.79 (0.73, 0.85) | 0.08 (0.07, 0.09) | 0.71 (0.65, 0.77) |  |
| 55 to <75 | 5.40 (4.52, 6.44) | 0.67 (0.59, 0.75) | 0.12 (0.11, 0.14) | 0.55 (0.47, 0.63) |  |
| 75 plus | 2.75 (2.03, 3.72) | 0.40 (0.30, 0.49) | 0.14 (0.12, 0.17) | 0.25 (0.15, 0.35) |  |
| **Gender** |  |  |  |  |  |
| Male | 8.69 (7.67, 9.85) | 0.86 (0.80, 0.93) | 0.10 (0.09, 0.11) | 0.76 (0.70, 0.83) | 0.41 |
| Female | 9.32 (8.37, 10.4) | 0.87 (0.82, 0.93) | 0.09 (0.09, 0.10) | 0.78 (0.73, 0.84) |  |
| **Ethnicity** |  |  |  |  |  |
| White | 8.28 (7.58, 9.05) | 0.86 (0.81, 0.90) | 0.10 (0.10, 0.11) | 0.75 (0.71, 0.80) | 0.009 |
| South Asian | 6.46 (4.38, 9.51) | 0.95 (0.70, 1.21) | 0.15 (0.11, 0.19) | 0.80 (0.55, 1.06) |  |
| Black | 14.7 (10.4, 21.0) | 1.08 (0.89, 1.27) | 0.07 (0.05, 0.10) | 1.01 (0.82, 1.20) |  |
| Other | 7.35 (3.68, 14.7) | 0.68 (0.44, 0.93) | 0.09 (0.04, 0.15) | 0.59 (0.34, 0.84) |  |
| Mixed | 11.8 (7.16, 19.3) | 0.94 (0.64, 1.23) | 0.08 (0.05, 0.11) | 0.86 (0.56, 1.15) |  |
| **Area-based deprivation** |  |  |  |  |  |
| 1 (least deprived) | 8.48 (6.72, 10.7) | 0.77 (0.66, 0.88) | 0.09 (0.07, 0.11) | 0.68 (0.57, 0.79) | 0.84 |
| 2 | 8.48 (6.92, 10.4) | 0.79 (0.70, 0.89) | 0.09 (0.08, 0.11) | 0.70 (0.60, 0.80) |  |
| 3 | 9.04 (7.67, 10.7) | 0.86 (0.78, 0.94) | 0.09 (0.08, 0.11) | 0.76 (0.68, 0.85) |  |
| 4 | 9.78 (8.19, 11.7) | 0.94 (0.83, 1.05) | 0.10 (0.08, 0.11) | 0.84 (0.73, 0.95) |  |
| 5 (most deprived) | 8.90 (7.55, 10.5) | 0.91 (0.83, 0.99) | 0.10 (0.09, 0.12) | 0.81 (0.73, 0.89) |  |
| **Urban-Rural practice** |  |  |  |  |  |
| urban | 9.22 (8.45, 10.1) | 0.89 (0.85, 0.94) | 0.10 (0.09, 0.10) | 0.80 (0.75, 0.84) | 0.15 |
| rural | 7.66 (6.05, 9.70) | 0.72 (0.62, 0.81) | 0.09 (0.07, 0.11) | 0.62 (0.53, 0.72) |  |
| **Calendar year** |  |  |  |  |  |
| 1998-1999 | 2.86 (1.08, 7.53) | 0.32 (0.13, 0.51) | 0.11 (0.03, 0.20) | 0.21 (0.001, 0.41) | 0.03 |
| 2000-2004 | 7.58 (5.97, 9.62) | 0.59 (0.51, 0.67) | 0.08 (0.06, 0.09) | 0.51 (0.43, 0.60) |  |
| 2005-2009 | 8.10 (6.89, 9.53) | 0.68 (0.62, 0.75) | 0.08 (0.07, 0.10) | 0.60 (0.53, 0.66) |  |
| 2010-2014 | 9.25 (8.01, 10.7) | 0.91 (0.83, 0.99) | 0.10 (0.09, 0.11) | 0.81 (0.73, 0.89) |  |
| 2015-2019 | 9.83 (8.50, 11.4) | 1.10 (1.01, 1.19) | 0.11 (0.10, 0.13) | 0.99 (0.90, 1.08) |  |

*includes 1998-1999 because index date is within coverage period but follow-up is sometimes earlier.

**SA3c Admitted patient care (all events)**

| **Covariate / level** | **Rate Ratio**  **(95% CI)** | **Rate per person-year in narcolepsy group** | **Rate per person-year in comparison group** | **Rate Difference per person-year (95% CI)** | **Interaction p-value (95% CI)** |
| --- | --- | --- | --- | --- | --- |
| **Age group** |  |  |  |  |  |
| <18 | 4.83 (3.40, 6.87) | 0.47 (0.33, 0.60) | 0.10 (0.08, 0.12) | 0.37 (0.23, 0.50) | <0.001 |
| 18 to <35 | 2.29 (1.98, 2.65) | 0.38 (0.33, 0.42) | 0.17 (0.15, 0.18) | 0.21 (0.17, 0.26) |  |
| 35 to <55 | 3.07 (2.31, 4.09) | 0.60 (0.45, 0.75) | 0.20 (0.17, 0.22) | 0.41 (0.26, 0.56) |  |
| 55 to <75 | 2.16 (1.72, 2.71) | 0.75 (0.67, 0.84) | 0.35 (0.28, 0.42) | 0.40 (0.29, 0.51) |  |
| 75 plus | 1.64 (1.23, 2.19) | 1.08 (0.89, 1.27) | 0.66 (0.51, 0.80) | 0.42 (0.18, 0.66) |  |
| **Gender** |  |  |  |  |  |
| Male | 2.19 (1.75, 2.73) | 0.53 (0.47, 0.58) | 0.24 (0.19, 0.29) | 0.29 (0.21, 0.36) | 0.16 |
| Female | 2.68 (2.27, 3.17) | 0.68 (0.58, 0.79) | 0.25 (0.24, 0.27) | 0.43 (0.32, 0.53) |  |
| **Ethnicity** |  |  |  |  |  |
| White | 2.35 (2.03, 2.73) | 0.63 (0.56, 0.70) | 0.27 (0.24, 0.30) | 0.36 (0.29, 0.43) | 0.68 |
| South Asian | 1.81 (1.22, 2.69) | 0.52 (0.36, 0.68) | 0.29 (0.21, 0.36) | 0.23 (0.06, 0.41) |  |
| Black | 1.78 (0.87, 3.62) | 0.66 (0.38, 0.93) | 0.37 (0.16, 0.58) | 0.29 (-0.06, 0.63) |  |
| Other | 1.82 (0.93, 3.53) | 0.33 (0.19, 0.47) | 0.18 (0.09, 0.28) | 0.15 (-0.02, 0.32) |  |
| Mixed | 2.17 (1.36, 3.48) | 0.51 (0.34, 0.68) | 0.24 (0.15, 0.32) | 0.28 (0.09, 0.46) |  |
| **Area-based deprivation** |  |  |  |  |  |
| 1 (least deprived) | 2.79 (1.32, 5.89) | 0.79 (0.38, 1.21) | 0.28 (0.13, 0.43) | 0.51 (0.07, 0.96) | 0.91 |
| 2 | 2.41 (2.01, 2.89) | 0.52 (0.44, 0.60) | 0.22 (0.20, 0.24) | 0.30 (0.22, 0.39) |  |
| 3 | 2.52 (2.21, 2.88) | 0.53 (0.47, 0.59) | 0.21 (0.20, 0.23) | 0.32 (0.26, 0.38) |  |
| 4 | 2.43 (1.85, 3.20) | 0.68 (0.59, 0.77) | 0.28 (0.21, 0.35) | 0.40 (0.29, 0.51) |  |
| 5 (most deprived) | 2.26 (1.87, 2.73) | 0.60 (0.52, 0.69) | 0.27 (0.23, 0.30) | 0.34 (0.24, 0.43) |  |
| **Urban-Rural practice** |  |  |  |  |  |
| urban | 2.48 (2.16, 2.85) | 0.61 (0.54, 0.68) | 0.25 (0.23, 0.27) | 0.37 (0.29, 0.44) | 0.79 |
| rural | 2.32 (1.46, 3.71) | 0.59 (0.49, 0.70) | 0.25 (0.15, 0.36) | 0.34 (0.18, 0.49) |  |
| **Calendar year** |  |  |  |  |  |
| 1998-1999 | 2.16 (1.60, 2.92) | 0.37 (0.27, 0.46) | 0.17 (0.14, 0.19) | 0.20 (0.10, 0.30) | 0.38 |
| 2000-2004 | 2.06 (1.54, 2.75) | 0.45 (0.39, 0.52) | 0.22 (0.17, 0.27) | 0.23 (0.15, 0.32) |  |
| 2005-2009 | 2.56 (2.09, 3.13) | 0.62 (0.53, 0.72) | 0.24 (0.21, 0.27) | 0.38 (0.28, 0.48) |  |
| 2010-2014 | 2.23 (1.69, 2.95) | 0.60 (0.52, 0.67) | 0.27 (0.20, 0.33) | 0.33 (0.23, 0.43) |  |
| 2015-2019 | 2.97 (2.21, 4.00) | 0.80 (0.58, 1.02) | 0.27 (0.23, 0.30) | 53.04 (31.24, 74.83) |  |

**SA3d Admitted patient care (possible-sleep related)**

| **Covariate / level** | **Rate Ratio (95% CI)** | **Rate per person-year in narcolepsy group** | **Rate per person-year in comparison group** | **Rate Difference per person-year (95% CI)** | **Interaction p-value (95% CI)** |
| --- | --- | --- | --- | --- | --- |
| **Age group** |  |  |  |  |  |
| <18 | 6.04 (3.97, 9.18) | 0.30 (0.22, 0.38) | 0.05 (0.03, 0.07) | 0.25 (0.17, 0.33) | <0.001 |
| 18 to <35 | 11.3 (9.22, 13.9) | 0.08 (0.07, 0.09) | 0.01 (0.01, 0.01) | 0.07 (0.06, 0.08) |  |
| 35 to <55 | 13.0 (9.85, 17.2) | 0.11 (0.09, 0.12) | 0.01 (0.01, 0.01) | 0.10 (0.08, 0.12) |  |
| 55 to <75 | 6.25 (4.91, 7.96) | 0.08 (0.06, 0.09) | 0.01 (0.01, 0.01) | 0.06 (0.05, 0.08) |  |
| 75 plus | 3.72 (2.03, 6.83) | 0.07 (0.03, 0.12) | 0.02 (0.02, 0.02) | 0.05 (0.01, 0.10) |  |
| **Gender** |  |  |  |  |  |
| Male | 7.12 (5.26, 9.65) | 0.12 (0.10, 0.14) | 0.02 (0.01, 0.02) | 0.10 (0.08, 0.13) | 0.26 |
| Female | 8.71 (7.32, 10.4) | 0.11 (0.10, 0.12) | 0.01 (0.01, 0.01) | 0.10 (0.08, 0.11) |  |
| **Ethnicity** |  |  |  |  |  |
| White | 7.79 (6.65, 9.12) | 0.12 (0.10, 0.13) | 0.01 (0.01, 0.02) | 0.10 (0.09, 0.12) | 0.32 |
| South Asian | 3.84 (1.21, 12.2) | 0.15 (0.05, 0.24) | 0.04 (<0.001, 0.08) | 0.11 (0.01, 0.21) |  |
| Black | 11.0 (7.50, 16.2) | 0.12 (0.09, 0.15) | 0.01 (0.01, 0.01) | 0.11 (0.08, 0.14) |  |
| Other | 6.91 (2.74, 17.4) | 0.07 (0.04, 0.11) | 0.01 (0.002, 0.02) | 0.06 (0.03, 0.10) |  |
| Mixed | 6.74 (3.35, 13.6) | 0.11 (0.06, 0.17) | 0.02 (0.01, 0.03) | 0.10 (0.04, 0.15) |  |
| **Area-based deprivation** |  |  |  |  |  |
| 1 (least deprived) | 7.98 (5.64, 11.3) | 0.11 (0.08, 0.14) | 0.01 (0.01, 0.02) | 0.10 (0.07, 0.13) | 0.41 |
| 2 | 8.48 (6.25, 11.5) | 0.09 (0.08, 0.11) | 0.01 (0.01, 0.01) | 0.08 (0.06, 0.10) |  |
| 3 | 8.64 (6.78, 11.0) | 0.11 (0.09, 0.14) | 0.01 (0.01, 0.02) | 0.10 (0.08, 0.12) |  |
| 4 | 9.71 (6.61, 14.2) | 0.14 (0.10, 0.18) | 0.01 (0.01, 0.02) | 0.13 (0.09, 0.17) |  |
| 5 (most deprived) | 5.61 (3.65, 8.62) | 0.11 (0.09, 0.12) | 0.02 (0.01, 0.03) | 0.09 (0.07, 0.11) |  |
| **Urban-Rural practice** |  |  |  |  |  |
| urban | 8.00 (6.51, 9.82) | 0.12 (0.11, 0.13) | 0.01 (0.01, 0.02) | 0.10 (0.09, 0.12) | 0.34 |
| rural | 6.51 (4.52, 9.39) | 0.09 (0.07, 0.11) | 0.01 (0.01, 0.02) | 0.07 (0.05, 0.09) |  |
| **Calendar year** |  |  |  |  |  |
| 1998-1999 | 9.09 (4.84, 17.1) | 0.10 (0.06, 0.14) | 0.01 (0.01, 0.02) | 0.09 (0.04, 0.13) | 0.28 |
| 2000-2004 | 6.65 (5.07, 8.72) | 0.06 (0.05, 0.08) | 0.01 (0.01, 0.01) | 0.05 (0.04, 0.07) |  |
| 2005-2009 | 5.62 (3.28, 9.62) | 0.10 (0.08, 0.11) | 0.02 (0.01, 0.03) | 0.08 (0.06, 0.10) |  |
| 2010-2014 | 9.22 (6.85, 12.4) | 0.13 (0.10, 0.17) | 0.01 (0.01, 0.02) | 0.12 (0.09, 0.15) |  |
| 2015-2019 | 8.68 (6.89, 10.9) | 0.15 (0.13, 0.17) | 0.02 (0.01, 0.02) | 0.13 (0.11, 0.15) |  |

**SA3e Accident & Emergency (all events)**

| **Covariate / level** | **Rate Ratio (95% CI)** | **Rate per person-year in narcolepsy group** | **Rate per person-year in comparison group** | **Rate Difference per person-year (95% CI)** | **Interaction p-value (95% CI)** |
| --- | --- | --- | --- | --- | --- |
| **Age group** |  |  |  |  |  |
| <18 | 1.55 (1.33, 1.81) | 0.41 (0.35, 0.46) | 0.26 (0.24, 0.28) | 0.15 (0.09, 0.20) | <0.001 |
| 18 to <35 | 1.97 (1.72, 2.26) | 0.43 (0.37, 0.48) | 0.22 (0.20, 0.23) | 0.21 (0.16, 0.26) |  |
| 35 to <55 | 2.56 (2.24, 2.94) | 0.47 (0.41, 0.52) | 0.18 (0.17, 0.20) | 0.29 (0.23, 0.34) |  |
| 55 to <75 | 3.01 (2.52, 3.60) | 0.44 (0.37, 0.51) | 0.15 (0.13, 0.16) | 0.29 (0.22, 0.36) |  |
| 75 plus | 2.05 (1.54, 2.73) | 0.69 (0.50, 0.88) | 0.34 (0.31, 0.36) | 0.35 (0.16, 0.54) |  |
| **Gender** |  |  |  |  |  |
| Male | 2.03 (1.82, 2.27) | 0.43 (0.38, 0.47) | 0.21 (0.20, 0.22) | 0.22 (0.17, 0.26) | 0.05 |
| Female | 2.38 (2.13, 2.65) | 0.49 (0.44, 0.54) | 0.21 (0.20, 0.22) | 0.28 (0.24, 0.33) |  |
| **Ethnicity** |  |  |  |  |  |
| White | 2.16 (1.98, 2.35) | 0.47 (0.43, 0.51) | 0.22 (0.21, 0.23) | 0.25 (0.22, 0.29) | 0.03 |
| South Asian | 1.85 (1.34, 2.56) | 0.49 (0.34, 0.64) | 0.27 (0.23, 0.30) | 0.23 (0.08, 0.38) |  |
| Black | 1.75 (1.32, 2.32) | 0.49 (0.38, 0.61) | 0.28 (0.24, 0.33) | 0.21 (0.09, 0.34) |  |
| Other | 0.79 (0.39, 1.60) | 0.17 (0.06, 0.29) | 0.22 (0.16, 0.27) | -0.05 (-0.17, 0.08) |  |
| Mixed | 1.71 (1.09, 2.70) | 0.42 (0.25, 0.58) | 0.24 (0.19, 0.30) | 0.17 (<-0.001, 0.35) |  |
| **Area-based deprivation** |  |  |  |  |  |
| 1 (least deprived) | 2.42 (1.75, 3.34) | 0.43 (0.30, 0.56) | 0.18 (0.16, 0.19) | 0.25 (0.12, 0.38) | 0.27 |
| 2 | 2.23 (1.86, 2.66) | 0.38 (0.32, 0.44) | 0.17 (0.16, 0.18) | 0.21 (0.15, 0.27) |  |
| 3 | 2.20 (1.90, 2.55) | 0.41 (0.35, 0.46) | 0.19 (0.17, 0.20) | 0.22 (0.17, 0.28) |  |
| 4 | 2.48 (2.10, 2.92) | 0.54 (0.46, 0.62) | 0.22 (0.20, 0.23) | 0.32 (0.24, 0.41) |  |
| 5 (most deprived) | 1.96 (1.71, 2.24) | 0.52 (0.46, 0.58) | 0.26 (0.25, 0.28) | 0.25 (0.19, 0.32) |  |
| **Urban-Rural practice** |  |  |  |  |  |
| urban | 2.17 (1.99, 2.36) | 0.47 (0.44, 0.51) | 0.22 (0.21, 0.23) | 0.25 (0.22, 0.29) | 0.13 |
| rural | 2.59 (2.09, 3.20) | 0.39 (0.32, 0.47) | 0.15 (0.14, 0.17) | 0.24 (0.16, 0.32) |  |
| **Calendar year*** |  |  |  |  |  |
| 1998-1999 | 1.01 (., .) | <0.001 (., .) | <0.001 (., .) | <0.001 (., .) | 0.64 |
| 2000-2004 | 2.06 (., .) | 0.06 (., .) | 0.03 (., .) | 0.03 (., .) |  |
| 2005-2009 | 2.16 (., .) | 0.29 (., .) | 0.14 (., .) | 0.16 (., .) |  |
| 2010-2014 | 2.17 (., .) | 0.56 (., .) | 0.26 (., .) | 0.30 (., .) |  |
| 2015-2019 | 2.30 (., .) | 0.70 (., .) | 0.30 (., .) | 0.39 (., .) |  |

*model did not converge properly

**SA3f Primary care (all events)**

| **Covariate / level** | **Rate Ratio (95% CI)** | **Rate per person-year in narcolepsy group** | **Rate per person-year in comparison group** | **Rate Difference per person-year (95% CI)** | **Interaction p-value (95% CI)** |
| --- | --- | --- | --- | --- | --- |
| **Age group** |  |  |  |  |  |
| <18 | 1.75 (1.60, 1.91) | 3.90 (3.60, 4.20) | 2.23 (2.13, 2.33) | 1.67 (1.35, 1.98) | <0.001 |
| 18 to <35 | 1.90 (1.77, 2.04) | 6.17 (5.78, 6.56) | 3.24 (3.13, 3.35) | 2.93 (2.53, 3.33) |  |
| 35 to <55 | 2.15 (2.00, 2.31) | 8.32 (7.79, 8.85) | 3.86 (3.74, 3.99) | 4.46 (3.91, 5.00) |  |
| 55 to <75 | 1.85 (1.68, 2.02) | 10.8 (9.87, 11.7) | 5.85 (5.65, 6.04) | 4.94 (4.00, 5.88) |  |
| 75 plus | 1.34 (1.21, 1.48) | 13.3 (12.2, 14.5) | 9.95 (9.48, 10.4) | 3.40 (2.14, 4.66) |  |
| **Gender** |  |  |  |  |  |
| Male | 1.91 (1.80, 2.03) | 7.52 (7.13, 7.91) | 3.93 (3.80, 4.06) | 3.59 (3.18, 3.99) | 0.22 |
| Female | 1.81 (1.71, 1.92) | 10.7 (10.1, 11.3) | 5.90 (5.76, 6.04) | 4.81 (4.22, 5.40) |  |
| **Ethnicity** |  |  |  |  |  |
| White | 1.81 (1.72, 1.89) | 9.38 (8.98, 9.78) | 5.19 (5.09, 5.30) | 4.18 (3.78, 4.59) | 0.14 |
| South Asian | 1.55 (1.24, 1.94) | 10.2 (8.06, 12.4) | 6.59 (6.06, 7.12) | 3.64 (1.42, 5.86) |  |
| Black | 1.56 (1.31, 1.85) | 8.67 (7.57, 9.78) | 5.57 (4.89, 6.25) | 3.10 (1.82, 4.38) |  |
| Other | 1.27 (0.86, 1.88) | 6.32 (4.28, 8.35) | 4.98 (3.84, 6.12) | 1.34 (-0.99, 3.66) |  |
| Mixed | 1.68 (1.21, 2.33) | 9.75 (7.06, 12.4) | 5.80 (4.76, 6.84) | 3.95 (1.08, 6.81) |  |
| **Area-based deprivation** |  |  |  |  |  |
| 1 (least deprived) | 1.80 (1.62, 2.00) | 9.03 (8.18, 9.88) | 5.01 (4.76, 5.27) | 4.02 (3.14, 4.90) | 0.94 |
| 2 | 1.87 (1.69, 2.06) | 9.28 (8.44, 10.1) | 4.97 (4.76, 5.19) | 4.30 (3.44, 5.17) |  |
| 3 | 1.90 (1.72, 2.10) | 9.51 (8.61, 10.4) | 5.00 (4.81, 5.19) | 4.50 (3.59, 5.42) |  |
| 4 | 1.81 (1.67, 1.97) | 8.89 (8.26, 9.51) | 4.90 (4.70, 5.11) | 3.98 (3.33, 4.64) |  |
| 5 (most deprived) | 1.86 (1.71, 2.02) | 9.28 (8.58, 9.98) | 4.99 (4.79, 5.19) | 4.29 (3.57, 5.01) |  |
| **Urban-Rural practice** |  |  |  |  |  |
| urban | 1.84 (1.76, 1.93) | 9.18 (8.79, 9.56) | 4.98 (4.87, 5.08) | 4.20 (3.80, 4.59) | 0.60 |
| rural | 1.90 (1.70, 2.14) | 9.48 (8.49, 10.5) | 4.98 (4.73, 5.23) | 4.50 (3.49, 5.52) |  |
| **Calendar year** |  |  |  |  |  |
| 1998-1999 | 1.68 (1.36, 2.09) | 5.53 (4.46, 6.60) | 3.28 (2.98, 3.59) | 2.25 (1.13, 3.36) | 0.14 |
| 2000-2004 | 1.69 (1.55, 1.85) | 6.86 (6.31, 7.41) | 4.05 (3.88, 4.22) | 2.81 (2.23, 3.38) |  |
| 2005-2009 | 1.87 (1.70, 2.05) | 9.43 (8.60, 10.3) | 5.05 (4.86, 5.24) | 4.38 (3.53, 5.22) |  |
| 2010-2014 | 1.86 (1.72, 2.02) | 10.2 (9.49, 10.9) | 5.48 (5.28, 5.69) | 4.74 (3.98, 5.49) |  |
| 2015-2019 | 1.97 (1.82, 2.13) | 10.9 (10.1, 11.7) | 5.54 (5.34, 5.74) | 5.37 (4.57, 6.17) |  |

**SA Tables 4a to 4b: Sex-stratified rate ratios, rates and rate differences for outpatient and admitted patient care events to specific consultant specialties**

**SA 4a: Outpatient**

| **Outcome** | **Sex** | **Rate Ratio (95% CI)** | **Rate per person-year in narcolepsy group  (95% CI)** | **Rate per person-year in comparison group  (95% CI)** | **Rate Difference per person-year (95% CI)** | **Interaction p-value (95% CI)** |
| --- | --- | --- | --- | --- | --- | --- |
| Respiratory Medicine | Male | 15.9 (12.3, 20.5) | 0.29 (0.25, 0.32) | 0.02 (0.01, 0.02) | 0.27 (0.23, 0.30) | 0.21 |
|  | Female | 20.0 (15.5, 25.9) | 0.29 (0.26, 0.32) | 0.01 (0.01, 0.02) | 0.28 (0.25, 0.31) |  |
| Neurology | Male | 11.7 (9.02, 15.2) | 0.22 (0.19, 0.24) | 0.02 (0.01, 0.02) | 0.20 (0.17, 0.22) | 0.26 |
|  | Female | 14.2 (11.5, 17.4) | 0.27 (0.25, 0.30) | 0.02 (0.02, 0.02) | 0.25 (0.23, 0.28) |  |
| Paediatrics | Male | 12.8 (9.43, 17.4) | 1.60 (1.32, 1.88) | 0.12 (0.09, 0.16) | 1.47 (1.19, 1.75) | 0.58 |
|  | Female | 11.4 (8.59, 15.1) | 1.50 (1.28, 1.73) | 0.13 (0.10, 0.16) | 1.37 (1.15, 1.60) |  |
| Ear Nose and Throat | Male | 2.19 (1.77, 2.71) | 0.10 (0.08, 0.12) | 0.05 (0.04, 0.05) | 0.05 (0.04, 0.07) | 0.39 |
|  | Female | 2.48 (2.05, 3.01) | 0.11 (0.09, 0.13) | 0.04 (0.04, 0.05) | 0.07 (0.05, 0.08) |  |
| Cardiothoracic Surgery | Male | 1.23 (0.69, 2.19) | 0.005 (0.002, 0.01) | 0.004 (0.003, 0.005) | <0.001 (-0.002, 0.003) | 0.65 |
|  | Female | 1.74 (0.43, 7.11) | 0.004 (<-0.001, 0.01) | 0.002 (<0.001, 0.003) | 0.002 (-0.003, 0.01) |  |
| General Surgery | Male | 1.98 (1.67, 2.34) | 0.12 (0.10, 0.14) | 0.06 (0.06, 0.07) | 0.06 (0.04, 0.08) | 0.60 |
|  | Female | 1.86 (1.58, 2.19) | 0.20 (0.17, 0.23) | 0.11 (0.10, 0.12) | 0.09 (0.06, 0.12) |  |
| Neurosurgery | Male | 3.59 (2.15, 5.99) | 0.02 (0.01, 0.02) | 0.005 (0.003, 0.01) | 0.01 (0.005, 0.02) | 0.25 |
|  | Female | 5.38 (3.37, 8.58) | 0.02 (0.02, 0.03) | 0.005 (0.003, 0.01) | 0.02 (0.01, 0.03) |  |
| Ophthalmology | Male | 1.59 (1.26, 2.02) | 0.18 (0.14, 0.21) | 0.11 (0.10, 0.12) | 0.07 (0.03, 0.10) | 0.59 |
|  | Female | 1.75 (1.37, 2.22) | 0.22 (0.17, 0.26) | 0.12 (0.11, 0.14) | 0.09 (0.04, 0.14) |  |
| Oral/Dentistry | Male | 2.04 (1.38, 3.01) | 0.09 (0.06, 0.12) | 0.04 (0.03, 0.05) | 0.04 (0.01, 0.07) | 0.83 |
|  | Female | 2.16 (1.61, 2.89) | 0.12 (0.09, 0.15) | 0.05 (0.05, 0.06) | 0.06 (0.03, 0.09) |  |
| Plastic Surgery | Male | 1.74 (1.14, 2.65) | 0.02 (0.02, 0.03) | 0.01 (0.01, 0.02) | 0.01 (0.001, 0.02) | 0.20 |
|  | Female | 2.72 (1.60, 4.64) | 0.05 (0.02, 0.07) | 0.02 (0.01, 0.02) | 0.03 (0.01, 0.05) |  |
| Trauma and Orthopaedics | Male | 1.67 (1.41, 1.97) | 0.20 (0.17, 0.23) | 0.12 (0.11, 0.13) | 0.08 (0.05, 0.11) | 0.22 |
|  | Female | 1.92 (1.65, 2.23) | 0.26 (0.22, 0.29) | 0.13 (0.12, 0.14) | 0.12 (0.09, 0.16) |  |
| Urology | Male | 1.67 (1.36, 2.05) | 0.11 (0.09, 0.13) | 0.07 (0.06, 0.08) | 0.05 (0.02, 0.07) | 0.004 |
|  | Female | 3.23 (2.16, 4.83) | 0.06 (0.04, 0.07) | 0.02 (0.01, 0.02) | 0.04 (0.02, 0.06) |  |
| Anaesthetics | Male | 3.64 (2.56, 5.17) | 0.03 (0.02, 0.04) | 0.01 (0.01, 0.01) | 0.02 (0.01, 0.03) | 0.04 |
|  | Female | 5.90 (4.38, 7.93) | 0.10 (0.07, 0.12) | 0.02 (0.01, 0.02) | 0.08 (0.06, 0.10) |  |
| Emergency Medicine | Male | 2.01 (1.29, 3.16) | 0.01 (0.004, 0.01) | 0.003 (0.002, 0.004) | 0.003 (<0.001, 0.01) | 0.15 |
|  | Female | 1.25 (0.79, 2.00) | 0.005 (0.003, 0.01) | 0.004 (0.003, 0.005) | 0.001 (-0.001, 0.003) |  |
| Intensive Care Medicine | Male | 4.49 (0.52, 39.0) | 0.002 (<0.001, 0.003) | <0.001 (<-0.001, 0.001) | 0.001 (<-0.001, 0.003) | 0.82 |
|  | Female | 5.94 (2.02, 17.4) | 0.002 (<0.001, 0.003) | <0.001 (<0.001, <0.001) | 0.001 (<0.001, 0.003) |  |
| Cardiology | Male | 1.96 (1.50, 2.57) | 0.09 (0.07, 0.12) | 0.05 (0.04, 0.05) | 0.05 (0.02, 0.07) | 0.04 |
|  | Female | 2.87 (2.24, 3.67) | 0.10 (0.08, 0.11) | 0.03 (0.03, 0.04) | 0.06 (0.04, 0.08) |  |
| Endocrinology and Diabetes | Male | 4.66 (2.73, 7.97) | 0.04 (0.02, 0.06) | 0.01 (0.01, 0.01) | 0.03 (0.02, 0.05) | 0.08 |
|  | Female | 2.58 (1.74, 3.81) | 0.04 (0.03, 0.06) | 0.02 (0.01, 0.02) | 0.03 (0.01, 0.04) |  |
| Renal Medicine | Male | 1.21 (0.55, 2.66) | 0.02 (0.01, 0.04) | 0.02 (0.01, 0.03) | 0.004 (-0.01, 0.02) | 0.94 |
|  | Female | 1.26 (0.60, 2.61) | 0.02 (0.01, 0.03) | 0.02 (0.01, 0.03) | 0.004 (-0.01, 0.02) |  |
| General Internal Medicine | Male | 3.60 (2.94, 4.40) | 0.23 (0.20, 0.26) | 0.06 (0.05, 0.07) | 0.17 (0.13, 0.20) | 0.71 |
|  | Female | 3.41 (2.81, 4.14) | 0.22 (0.19, 0.25) | 0.06 (0.06, 0.07) | 0.16 (0.12, 0.19) |  |
| General Medical Practice | Male | 0.28 (0.06, 1.26) | <0.001 (<-0.001, 0.001) | 0.002 (<-0.001, 0.004) | -0.001 (-0.003, <0.001) | 0.004 |
|  | Female | 7.83 (1.44, 42.5) | 0.01 (-0.004, 0.02) | <0.001 (<0.001, 0.002) | 0.01 (-0.01, 0.02) |  |
| Geriatric Medicine | Male | 2.16 (1.31, 3.55) | 0.06 (0.04, 0.08) | 0.03 (0.02, 0.04) | 0.03 (0.01, 0.06) | 0.17 |
|  | Female | 3.42 (2.22, 5.27) | 0.09 (0.06, 0.12) | 0.03 (0.02, 0.03) | 0.06 (0.03, 0.10) |  |
| Audio Vestibular Medicine | Male | 2.04 (1.06, 3.92) | 0.01 (0.01, 0.02) | 0.01 (0.004, 0.01) | 0.01 (-0.001, 0.01) | 0.71 |
|  | Female | 2.43 (1.25, 4.69) | 0.01 (0.005, 0.02) | 0.005 (0.003, 0.01) | 0.01 (<-0.001, 0.01) |  |
| Clinical Haematology | Male | 1.03 (0.57, 1.85) | 0.06 (0.03, 0.09) | 0.06 (0.04, 0.08) | 0.002 (-0.03, 0.04) | 0.07 |
|  | Female | 2.12 (1.25, 3.60) | 0.07 (0.04, 0.10) | 0.03 (0.02, 0.04) | 0.04 (0.005, 0.07) |  |
| Clinical Immunology | Male | 2.17 (0.46, 10.2) | <0.001 (<-0.001, 0.002) | <0.001 (<-0.001, <0.001) | <0.001 (<-0.001, 0.002) | 0.78 |
|  | Female | 2.83 (0.95, 8.38) | 0.005 (<0.001, 0.01) | 0.002 (<0.001, 0.003) | 0.003 (-0.001, 0.01) |  |
| Clinical Pharmacology | Male | 0.75 (0.07, 8.50) | <0.001 (<-0.001, <0.001) | <0.001 (<-0.001, <0.001) | <-0.001 (<-0.001, <0.001) | 0.67 |
|  | Female | 1.60 (0.13, 19.9) | 0.003 (-0.003, 0.01) | 0.002 (-0.001, 0.01) | 0.001 (-0.01, 0.01) |  |
| Clinical Physiology | Male | 3.01 (1.36, 6.67) | 0.003 (<0.001, 0.005) | <0.001 (<0.001, 0.001) | 0.002 (<-0.001, 0.004) | 0.48 |
|  | Female | 2.01 (0.91, 4.47) | 0.003 (<0.001, 0.01) | 0.002 (0.001, 0.002) | 0.002 (<-0.001, 0.004) |  |
| Dermatology | Male | 1.23 (0.83, 1.81) | 0.06 (0.04, 0.08) | 0.05 (0.04, 0.06) | 0.01 (-0.01, 0.03) | 0.19 |
|  | Female | 1.70 (1.27, 2.28) | 0.10 (0.07, 0.12) | 0.06 (0.05, 0.07) | 0.04 (0.01, 0.07) |  |
| Gastroenterology | Male | 1.56 (1.06, 2.29) | 0.03 (0.02, 0.04) | 0.02 (0.02, 0.03) | 0.01 (<0.001, 0.02) | 0.01 |
|  | Female | 2.92 (2.17, 3.93) | 0.05 (0.04, 0.07) | 0.02 (0.02, 0.02) | 0.04 (0.02, 0.05) |  |
| Infectious Diseases | Male | 5.76 (1.47, 22.6) | 0.01 (-0.001, 0.01) | <0.001 (<0.001, 0.001) | 0.004 (-0.002, 0.01) | 0.86 |
|  | Female | 4.67 (0.76, 28.8) | 0.01 (-0.004, 0.02) | 0.001 (<0.001, 0.003) | 0.01 (-0.005, 0.02) |  |
| Medical Oncology | Male | 0.87 (0.44, 1.75) | 0.01 (0.01, 0.02) | 0.02 (0.01, 0.02) | -0.002 (-0.01, 0.01) | 0.15 |
|  | Female | 1.86 (0.88, 3.93) | 0.02 (0.01, 0.03) | 0.01 (0.01, 0.02) | 0.01 (-0.004, 0.02) |  |
| Palliative Medicine | Male | 0.93 (0.11, 8.16) | 0.003 (<-0.001, 0.01) | 0.004 (-0.002, 0.01) | <-0.001 (-0.01, 0.01) | 0.010 |
|  | Female | 49.8 (5.61, 442) | 0.03 (-0.02, 0.07) | <0.001 (<-0.001, 0.001) | 0.03 (-0.02, 0.07) |  |
| Psychiatry | Male | 4.71 (2.16, 10.3) | 0.20 (0.11, 0.29) | 0.04 (0.02, 0.07) | 0.16 (0.06, 0.25) | 0.67 |
|  | Female | 3.91 (2.83, 5.42) | 0.15 (0.11, 0.18) | 0.04 (0.03, 0.05) | 0.11 (0.08, 0.14) |  |
| Rehabilitation Medicine | Male | 8.25 (2.62, 26.0) | 0.03 (-0.004, 0.07) | 0.004 (0.002, 0.01) | 0.03 (-0.01, 0.07) | 0.03 |
|  | Female | 1.89 (0.87, 4.10) | 0.01 (0.01, 0.02) | 0.01 (0.003, 0.01) | 0.01 (-0.001, 0.01) |  |
| Rheumatology | Male | 2.06 (1.26, 3.37) | 0.03 (0.02, 0.04) | 0.02 (0.01, 0.02) | 0.02 (0.003, 0.03) | 0.80 |
|  | Female | 1.90 (1.30, 2.79) | 0.08 (0.06, 0.11) | 0.04 (0.03, 0.05) | 0.04 (0.01, 0.07) |  |
| Non-clinical/hospital consultant led | Male | 1.91 (1.37, 2.66) | 0.11 (0.08, 0.15) | 0.06 (0.05, 0.07) | 0.05 (0.02, 0.09) | 0.81 |
|  | Female | 2.01 (1.53, 2.64) | 0.16 (0.12, 0.19) | 0.08 (0.07, 0.09) | 0.08 (0.04, 0.12) |  |

**SA4b Admitted Patient Care**

| **Outcome** | **Sex** | **Rate Ratio (95% CI)** | **Rate per person-year in narcolepsy group  (95% CI)** | **Rate per person-year in comparison group  (95% CI)** | **Rate Difference per person-year (95% CI)** | **Interaction p-value (95% CI)** |
| --- | --- | --- | --- | --- | --- | --- |
| Respiratory Medicine | Male | 14.6 (11.1, 19.2) | 0.05 (0.04, 0.06) | 0.003 (0.003, 0.004) | 0.04 (0.04, 0.05) | 0.65 |
|  | Female | 13.3 (10.1, 17.6) | 0.04 (0.04, 0.05) | 0.003 (0.003, 0.004) | 0.04 (0.03, 0.05) |  |
| Neurology | Male | 10.4 (5.20, 20.9) | 0.02 (0.01, 0.02) | 0.002 (<0.001, 0.003) | 0.02 (0.01, 0.02) | 0.02 |
|  | Female | 28.5 (17.2, 47.2) | 0.03 (0.02, 0.03) | <0.001 (<0.001, 0.001) | 0.02 (0.02, 0.03) |  |
| Paediatrics | Male | 6.18 (2.98, 12.8) | 0.29 (0.17, 0.41) | 0.05 (0.02, 0.07) | 0.24 (0.12, 0.36) | 0.73 |
|  | Female | 7.20 (4.63, 11.2) | 0.22 (0.16, 0.27) | 0.03 (0.02, 0.04) | 0.19 (0.13, 0.24) |  |
| Ear Nose and Throat | Male | 2.64 (1.94, 3.60) | 0.02 (0.01, 0.02) | 0.01 (0.005, 0.01) | 0.01 (0.01, 0.01) | 0.88 |
|  | Female | 2.56 (1.97, 3.32) | 0.01 (0.01, 0.02) | 0.005 (0.004, 0.01) | 0.01 (0.005, 0.01) |  |
| Cardiothoracic Surgery | Male | 1.54 (0.92, 2.57) | 0.003 (0.001, 0.004) | 0.002 (0.001, 0.002) | <0.001 (<-0.001, 0.002) | 0.36 |
|  | Female | 2.66 (0.92, 7.65) | 0.002 (<0.001, 0.004) | <0.001 (<0.001, 0.001) | 0.001 (<-0.001, 0.003) |  |
| General Surgery | Male | 2.05 (1.77, 2.38) | 0.06 (0.05, 0.07) | 0.03 (0.03, 0.03) | 0.03 (0.02, 0.04) | 0.59 |
|  | Female | 2.18 (1.87, 2.54) | 0.07 (0.06, 0.08) | 0.03 (0.03, 0.03) | 0.04 (0.03, 0.05) |  |
| Neurosurgery | Male | 5.48 (3.25, 9.26) | 0.01 (0.004, 0.01) | 0.001 (<0.001, 0.002) | 0.01 (0.003, 0.01) | 0.40 |
|  | Female | 3.94 (2.24, 6.91) | 0.01 (0.004, 0.01) | 0.002 (0.001, 0.002) | 0.005 (0.002, 0.01) |  |
| Ophthalmology | Male | 1.22 (0.88, 1.70) | 0.01 (0.01, 0.02) | 0.01 (0.01, 0.01) | 0.002 (-0.002, 0.01) | 0.52 |
|  | Female | 1.40 (1.08, 1.83) | 0.01 (0.01, 0.02) | 0.01 (0.01, 0.01) | 0.004 (<0.001, 0.01) |  |
| Oral/Dentistry | Male | 1.87 (1.19, 2.95) | 0.01 (0.01, 0.01) | 0.005 (0.004, 0.01) | 0.004 (<0.001, 0.01) | 0.73 |
|  | Female | 2.06 (1.54, 2.75) | 0.01 (0.01, 0.01) | 0.005 (0.004, 0.01) | 0.005 (0.002, 0.01) |  |
| Plastic Surgery | Male | 1.48 (0.99, 2.22) | 0.005 (0.003, 0.01) | 0.003 (0.003, 0.004) | 0.002 (<-0.001, 0.003) | 0.28 |
|  | Female | 2.04 (1.35, 3.09) | 0.01 (0.01, 0.01) | 0.004 (0.003, 0.005) | 0.004 (0.001, 0.01) |  |
| Trauma and Orthopaedics | Male | 1.65 (1.33, 2.05) | 0.03 (0.02, 0.03) | 0.02 (0.02, 0.02) | 0.01 (0.01, 0.02) | 0.14 |
|  | Female | 2.04 (1.69, 2.46) | 0.04 (0.03, 0.05) | 0.02 (0.02, 0.02) | 0.02 (0.01, 0.03) |  |
| Urology | Male | 1.82 (1.32, 2.51) | 0.04 (0.03, 0.05) | 0.02 (0.02, 0.02) | 0.02 (0.01, 0.03) | 0.002 |
|  | Female | 4.61 (2.87, 7.41) | 0.03 (0.01, 0.04) | 0.01 (0.004, 0.01) | 0.02 (0.01, 0.03) |  |
| Anaesthetics | Male | 4.15 (2.28, 7.55) | 0.01 (0.005, 0.01) | 0.002 (0.001, 0.003) | 0.01 (0.002, 0.01) | 0.93 |
|  | Female | 4.30 (2.38, 7.78) | 0.02 (0.01, 0.03) | 0.005 (0.003, 0.01) | 0.02 (0.01, 0.02) |  |
| Emergency Medicine | Male | 2.40 (1.80, 3.21) | 0.02 (0.02, 0.03) | 0.01 (0.01, 0.01) | 0.01 (0.01, 0.02) | 0.18 |
|  | Female | 3.30 (2.29, 4.75) | 0.03 (0.02, 0.04) | 0.01 (0.01, 0.01) | 0.02 (0.01, 0.03) |  |
| Intensive Care Medicine | Male | 4.98 (2.31, 10.8) | 0.002 (<0.001, 0.002) | <0.001 (<0.001, <0.001) | 0.001 (<0.001, 0.002) | 0.17 |
|  | Female | 11.0 (4.73, 25.6) | 0.002 (<0.001, 0.004) | <0.001 (<0.001, <0.001) | 0.002 (<0.001, 0.004) |  |
| Cardiology | Male | 1.80 (1.37, 2.37) | 0.02 (0.01, 0.02) | 0.01 (0.01, 0.01) | 0.01 (0.004, 0.01) | 0.003 |
|  | Female | 3.30 (2.48, 4.38) | 0.02 (0.02, 0.02) | 0.01 (0.01, 0.01) | 0.01 (0.01, 0.02) |  |
| Endocrinology and Diabetes | Male | 5.94 (3.60, 9.81) | 0.01 (0.005, 0.01) | 0.001 (<0.001, 0.002) | 0.01 (0.004, 0.01) | 0.73 |
|  | Female | 5.27 (3.31, 8.40) | 0.01 (0.004, 0.01) | 0.001 (0.001, 0.002) | 0.01 (0.003, 0.01) |  |
| Renal Medicine | Male | 0.31 (0.05, 2.06) | 0.01 (-0.01, 0.03) | 0.04 (-0.01, 0.08) | -0.03 (-0.08, 0.02) | 0.05 |
|  | Female | 4.60 (0.67, 31.7) | 0.06 (-0.03, 0.15) | 0.01 (-0.001, 0.03) | 0.04 (-0.05, 0.13) |  |
| Acute Internal Medicine | Male | 3.65 (1.80, 7.38) | 0.003 (0.001, 0.005) | <0.001 (<0.001, 0.001) | 0.002 (<0.001, 0.004) | 0.36 |
|  | Female | 6.08 (2.62, 14.1) | 0.004 (<0.001, 0.01) | <0.001 (<0.001, <0.001) | 0.003 (<0.001, 0.01) |  |
| General Internal Medicine | Male | 2.73 (2.24, 3.33) | 0.11 (0.09, 0.12) | 0.04 (0.03, 0.04) | 0.07 (0.05, 0.08) | 0.09 |
|  | Female | 3.40 (2.89, 4.01) | 0.12 (0.11, 0.14) | 0.04 (0.03, 0.04) | 0.09 (0.07, 0.10) |  |
| Geriatric Medicine | Male | 2.80 (1.99, 3.93) | 0.07 (0.05, 0.09) | 0.02 (0.02, 0.03) | 0.04 (0.02, 0.07) | 0.58 |
|  | Female | 3.21 (2.29, 4.51) | 0.07 (0.05, 0.09) | 0.02 (0.02, 0.02) | 0.05 (0.03, 0.07) |  |
| Clinical Haematology | Male | 0.92 (0.21, 4.02) | 0.01 (-0.004, 0.03) | 0.01 (0.01, 0.02) | -0.001 (-0.02, 0.02) | 0.17 |
|  | Female | 3.27 (1.15, 9.31) | 0.02 (0.002, 0.03) | 0.005 (0.002, 0.01) | 0.01 (-0.003, 0.02) |  |
| Dermatology | Male | 1.58 (0.87, 2.87) | 0.003 (0.002, 0.01) | 0.002 (0.002, 0.003) | 0.001 (<-0.001, 0.003) | 0.69 |
|  | Female | 1.89 (0.98, 3.64) | 0.003 (0.001, 0.01) | 0.002 (0.001, 0.002) | 0.002 (<-0.001, 0.004) |  |
| Gastroenterology | Male | 1.86 (1.43, 2.42) | 0.02 (0.02, 0.03) | 0.01 (0.01, 0.01) | 0.01 (0.01, 0.01) | 0.03 |
|  | Female | 2.81 (2.13, 3.70) | 0.03 (0.03, 0.04) | 0.01 (0.01, 0.01) | 0.02 (0.01, 0.03) |  |
| Infectious Diseases | Male | 3.82 (1.84, 7.94) | 0.002 (<0.001, 0.003) | <0.001 (<0.001, <0.001) | 0.001 (<0.001, 0.002) | 0.47 |
|  | Female | 2.64 (1.33, 5.26) | 0.001 (<0.001, 0.002) | <0.001 (<0.001, <0.001) | <0.001 (<0.001, 0.002) |  |
| Medical Oncology | Male | 1.25 (0.39, 3.97) | 0.01 (<-0.001, 0.01) | 0.005 (0.002, 0.01) | 0.001 (-0.01, 0.01) | 0.75 |
|  | Female | 1.63 (0.52, 5.07) | 0.01 (0.003, 0.02) | 0.01 (0.001, 0.02) | 0.01 (-0.01, 0.02) |  |
| Psychiatry | Male | 5.28 (1.76, 15.8) | 0.01 (0.005, 0.02) | 0.003 (<0.001, 0.01) | 0.01 (0.001, 0.02) | 0.54 |
|  | Female | 3.66 (2.35, 5.71) | 0.01 (0.01, 0.01) | 0.002 (0.001, 0.003) | 0.01 (0.003, 0.01) |  |
| Rehabilitation Medicine | Male | 4.26 (1.72, 10.5) | 0.001 (<0.001, 0.002) | <0.001 (<0.001, <0.001) | <0.001 (<0.001, 0.002) | 0.40 |
|  | Female | 9.52 (1.87, 48.4) | 0.003 (-0.002, 0.01) | <0.001 (<0.001, <0.001) | 0.003 (-0.002, 0.01) |  |
| Rheumatology | Male | 0.99 (0.39, 2.54) | 0.002 (<0.001, 0.003) | 0.002 (<0.001, 0.004) | <-0.001 (-0.002, 0.002) | 0.01 |
|  | Female | 4.79 (2.11, 10.9) | 0.02 (0.01, 0.03) | 0.004 (0.002, 0.01) | 0.01 (0.002, 0.03) |  |
| Non-clinical/hospital consultant led | Male | 3.57 (1.05, 12.1) | 0.02 (-0.003, 0.04) | 0.01 (0.003, 0.01) | 0.01 (-0.01, 0.04) | 0.12 |
|  | Female | 1.21 (0.70, 2.11) | 0.01 (0.01, 0.02) | 0.01 (0.01, 0.01) | 0.002 (-0.004, 0.01) |  |

**SA Tables 5a to 5b: Age-stratified rate ratios and rate differences for outpatient and admitted patient care events to specific consultant specialties**

**SA 5a: Outpatient**

| **Outcome** | **Age group** | **Rate Ratio (95% CI)** | **Rate per person-year in narcolepsy group  (95% CI)** | **Rate per person-year in comparison group  (95% CI)** | **Rate Difference per person-year (95% CI)** | **Interaction p-value (95% CI)** |
| --- | --- | --- | --- | --- | --- | --- |
| Respiratory Medicine | <18 | 22.3 (6.23, 80.1) | 0.12 (0.08, 0.16) | 0.01 (-0.001, 0.01) | 0.12 (0.08, 0.16) | <0.001 |
|  | 18 to <35 | 57.5 (35.8, 92.3) | 0.36 (0.31, 0.40) | 0.01 (0.003, 0.01) | 0.35 (0.31, 0.40) |  |
|  | 35 to <55 | 19.8 (14.6, 26.7) | 0.34 (0.30, 0.39) | 0.02 (0.01, 0.02) | 0.33 (0.28, 0.37) |  |
|  | 55 to <75 | 9.83 (7.15, 13.5) | 0.27 (0.22, 0.32) | 0.03 (0.02, 0.03) | 0.24 (0.19, 0.29) |  |
|  | 75 plus | 3.56 (2.09, 6.06) | 0.12 (0.07, 0.18) | 0.03 (0.02, 0.05) | 0.09 (0.03, 0.14) |  |
| Neurology | <18 | 40.1 (20.6, 78.2) | 0.11 (0.08, 0.14) | 0.003 (0.001, 0.004) | 0.11 (0.08, 0.14) | <0.001 |
|  | 18 to <35 | 21.5 (15.6, 29.5) | 0.29 (0.26, 0.32) | 0.01 (0.01, 0.02) | 0.28 (0.24, 0.31) |  |
|  | 35 to <55 | 16.0 (12.4, 20.7) | 0.30 (0.27, 0.34) | 0.02 (0.01, 0.02) | 0.28 (0.25, 0.32) |  |
|  | 55 to <75 | 6.39 (4.57, 8.93) | 0.22 (0.18, 0.26) | 0.03 (0.02, 0.04) | 0.19 (0.15, 0.22) |  |
|  | 75 plus | 5.34 (3.23, 8.81) | 0.14 (0.08, 0.19) | 0.03 (0.02, 0.03) | 0.11 (0.06, 0.17) |  |
| Ear Nose and Throat | <18 | 1.62 (1.15, 2.30) | 0.07 (0.05, 0.09) | 0.04 (0.03, 0.05) | 0.03 (0.01, 0.05) | 0.04 |
|  | 18 to <35 | 2.70 (2.00, 3.63) | 0.07 (0.05, 0.08) | 0.02 (0.02, 0.03) | 0.04 (0.03, 0.06) |  |
|  | 35 to <55 | 2.70 (2.15, 3.40) | 0.11 (0.09, 0.13) | 0.04 (0.03, 0.05) | 0.07 (0.05, 0.09) |  |
|  | 55 to <75 | 2.50 (1.86, 3.35) | 0.15 (0.12, 0.19) | 0.06 (0.05, 0.07) | 0.09 (0.05, 0.13) |  |
|  | 75 plus | 1.49 (0.91, 2.44) | 0.12 (0.07, 0.17) | 0.08 (0.06, 0.10) | 0.04 (-0.01, 0.09) |  |
| General Surgery | <18 | 1.27 (0.72, 2.24) | 0.02 (0.01, 0.03) | 0.02 (0.01, 0.02) | 0.004 (-0.01, 0.01) | 0.02 |
|  | 18 to <35 | 2.37 (1.81, 3.11) | 0.07 (0.06, 0.09) | 0.03 (0.03, 0.04) | 0.04 (0.02, 0.06) |  |
|  | 35 to <55 | 2.21 (1.82, 2.69) | 0.20 (0.16, 0.23) | 0.09 (0.08, 0.10) | 0.11 (0.07, 0.14) |  |
|  | 55 to <75 | 1.53 (1.25, 1.88) | 0.23 (0.19, 0.27) | 0.15 (0.13, 0.17) | 0.08 (0.04, 0.12) |  |
|  | 75 plus | 1.72 (1.28, 2.32) | 0.26 (0.20, 0.33) | 0.15 (0.13, 0.18) | 0.11 (0.04, 0.18) |  |
| Ophthalmology | <18 | 1.47 (0.87, 2.50) | 0.08 (0.05, 0.12) | 0.06 (0.04, 0.07) | 0.03 (-0.01, 0.07) | 0.01 |
|  | 18 to <35 | 2.23 (1.49, 3.34) | 0.05 (0.03, 0.07) | 0.02 (0.02, 0.03) | 0.03 (0.01, 0.05) |  |
|  | 35 to <55 | 2.07 (1.54, 2.78) | 0.12 (0.09, 0.15) | 0.06 (0.05, 0.07) | 0.06 (0.03, 0.09) |  |
|  | 55 to <75 | 1.92 (1.40, 2.64) | 0.32 (0.23, 0.41) | 0.17 (0.14, 0.19) | 0.15 (0.06, 0.25) |  |
|  | 75 plus | 1.19 (0.93, 1.53) | 0.59 (0.47, 0.72) | 0.50 (0.44, 0.56) | 0.10 (-0.05, 0.24) |  |
| Oral/Dentistry | <18 | 1.48 (0.85, 2.59) | 0.15 (0.07, 0.22) | 0.10 (0.08, 0.13) | 0.05 (-0.03, 0.13) | 0.09 |
|  | 18 to <35 | 2.38 (1.40, 4.05) | 0.10 (0.05, 0.15) | 0.04 (0.03, 0.05) | 0.06 (0.01, 0.11) |  |
|  | 35 to <55 | 3.05 (2.18, 4.28) | 0.10 (0.07, 0.13) | 0.03 (0.03, 0.04) | 0.07 (0.04, 0.10) |  |
|  | 55 to <75 | 1.80 (1.15, 2.80) | 0.10 (0.06, 0.13) | 0.05 (0.04, 0.07) | 0.04 (0.005, 0.08) |  |
|  | 75 plus | 1.31 (0.60, 2.86) | 0.06 (0.02, 0.10) | 0.04 (0.03, 0.06) | 0.01 (-0.03, 0.06) |  |
| Plastic Surgery | <18 | 1.34 (0.51, 3.54) | 0.02 (0.002, 0.03) | 0.01 (0.01, 0.02) | 0.005 (-0.01, 0.02) | 0.23 |
|  | 18 to <35 | 2.37 (1.33, 4.21) | 0.02 (0.01, 0.03) | 0.01 (0.01, 0.01) | 0.01 (0.002, 0.02) |  |
|  | 35 to <55 | 2.89 (1.70, 4.92) | 0.04 (0.02, 0.05) | 0.01 (0.01, 0.02) | 0.02 (0.01, 0.04) |  |
|  | 55 to <75 | 3.20 (1.22, 8.39) | 0.06 (0.005, 0.11) | 0.02 (0.01, 0.02) | 0.04 (-0.01, 0.09) |  |
|  | 75 plus | 1.10 (0.49, 2.47) | 0.05 (0.01, 0.08) | 0.04 (0.03, 0.06) | 0.004 (-0.03, 0.04) |  |
| Trauma and Orthopaedics | <18 | 1.43 (1.03, 1.97) | 0.14 (0.10, 0.18) | 0.10 (0.09, 0.11) | 0.04 (<-0.001, 0.09) | 0.06 |
|  | 18 to <35 | 1.54 (1.21, 1.94) | 0.11 (0.08, 0.13) | 0.07 (0.06, 0.08) | 0.04 (0.01, 0.06) |  |
|  | 35 to <55 | 2.15 (1.80, 2.58) | 0.22 (0.19, 0.26) | 0.10 (0.09, 0.11) | 0.12 (0.09, 0.15) |  |
|  | 55 to <75 | 1.88 (1.51, 2.34) | 0.38 (0.30, 0.45) | 0.20 (0.18, 0.22) | 0.18 (0.10, 0.25) |  |
|  | 75 plus | 1.51 (1.13, 2.00) | 0.33 (0.24, 0.41) | 0.22 (0.19, 0.24) | 0.11 (0.02, 0.20) |  |
| Urology | <18 | 1.63 (0.66, 4.01) | 0.01 (0.002, 0.01) | 0.01 (0.003, 0.01) | 0.003 (-0.004, 0.01) | 0.03 |
|  | 18 to <35 | 3.01 (1.85, 4.88) | 0.03 (0.02, 0.04) | 0.01 (0.01, 0.01) | 0.02 (0.01, 0.03) |  |
|  | 35 to <55 | 2.96 (1.96, 4.45) | 0.08 (0.06, 0.10) | 0.03 (0.02, 0.03) | 0.05 (0.03, 0.07) |  |
|  | 55 to <75 | 1.69 (1.28, 2.21) | 0.12 (0.09, 0.15) | 0.07 (0.06, 0.08) | 0.05 (0.02, 0.08) |  |
|  | 75 plus | 1.48 (1.03, 2.14) | 0.19 (0.13, 0.25) | 0.13 (0.11, 0.15) | 0.06 (-0.003, 0.13) |  |
| Anaesthetics | <18 | 7.04 (2.70, 18.4) | 0.02 (0.005, 0.03) | 0.002 (<0.001, 0.004) | 0.01 (0.003, 0.02) | 0.07 |
|  | 18 to <35 | 6.57 (3.81, 11.3) | 0.05 (0.03, 0.07) | 0.01 (0.005, 0.01) | 0.04 (0.02, 0.06) |  |
|  | 35 to <55 | 6.35 (4.32, 9.34) | 0.09 (0.07, 0.12) | 0.01 (0.01, 0.02) | 0.08 (0.05, 0.10) |  |
|  | 55 to <75 | 3.94 (2.61, 5.94) | 0.07 (0.05, 0.10) | 0.02 (0.01, 0.02) | 0.05 (0.03, 0.08) |  |
|  | 75 plus | 2.42 (1.25, 4.68) | 0.05 (0.02, 0.08) | 0.02 (0.01, 0.03) | 0.03 (<0.001, 0.06) |  |
| Cardiology | <18 | 5.57 (2.26, 13.7) | 0.04 (0.01, 0.07) | 0.01 (0.004, 0.01) | 0.03 (<0.001, 0.06) | 0.007 |
|  | 18 to <35 | 2.28 (1.47, 3.54) | 0.03 (0.02, 0.03) | 0.01 (0.01, 0.01) | 0.01 (0.005, 0.02) |  |
|  | 35 to <55 | 2.59 (1.84, 3.64) | 0.07 (0.05, 0.09) | 0.03 (0.02, 0.03) | 0.04 (0.02, 0.06) |  |
|  | 55 to <75 | 2.71 (2.00, 3.67) | 0.18 (0.14, 0.23) | 0.07 (0.06, 0.08) | 0.12 (0.07, 0.16) |  |
|  | 75 plus | 1.36 (0.96, 1.92) | 0.18 (0.12, 0.23) | 0.13 (0.11, 0.15) | 0.05 (-0.01, 0.10) |  |
| Endocrinology and Diabetes | <18 | 18.7 (5.77, 60.7) | 0.04 (0.01, 0.07) | 0.002 (<0.001, 0.005) | 0.04 (0.01, 0.07) | 0.01 |
|  | 18 to <35 | 2.44 (1.27, 4.68) | 0.02 (0.01, 0.04) | 0.01 (0.01, 0.01) | 0.01 (0.002, 0.03) |  |
|  | 35 to <55 | 2.41 (1.56, 3.73) | 0.05 (0.03, 0.06) | 0.02 (0.01, 0.03) | 0.03 (0.01, 0.04) |  |
|  | 55 to <75 | 4.43 (2.38, 8.26) | 0.07 (0.04, 0.10) | 0.02 (0.01, 0.02) | 0.05 (0.02, 0.09) |  |
|  | 75 plus | 2.38 (0.86, 6.59) | 0.02 (0.003, 0.04) | 0.01 (0.005, 0.02) | 0.01 (-0.01, 0.03) |  |
| General Internal Medicine | <18 | 7.60 (3.24, 17.8) | 0.03 (0.01, 0.04) | 0.004 (0.001, 0.01) | 0.02 (0.01, 0.04) | <0.001 |
|  | 18 to <35 | 3.87 (2.72, 5.52) | 0.14 (0.11, 0.17) | 0.04 (0.03, 0.05) | 0.10 (0.07, 0.14) |  |
|  | 35 to <55 | 4.74 (3.69, 6.09) | 0.26 (0.21, 0.30) | 0.05 (0.04, 0.06) | 0.20 (0.16, 0.25) |  |
|  | 55 to <75 | 2.85 (2.27, 3.58) | 0.31 (0.25, 0.37) | 0.11 (0.09, 0.12) | 0.20 (0.14, 0.26) |  |
|  | 75 plus | 2.23 (1.63, 3.04) | 0.31 (0.23, 0.40) | 0.14 (0.12, 0.17) | 0.17 (0.09, 0.26) |  |
| Clinical Haematology | <18 | 3.04 (1.08, 8.50) | 0.02 (0.01, 0.03) | 0.01 (0.002, 0.01) | 0.01 (-0.002, 0.03) | 0.31 |
|  | 18 to <35 | 1.94 (0.54, 6.95) | 0.03 (-0.001, 0.05) | 0.01 (0.004, 0.02) | 0.01 (-0.02, 0.04) |  |
|  | 35 to <55 | 2.19 (1.08, 4.43) | 0.05 (0.02, 0.08) | 0.02 (0.01, 0.03) | 0.03 (-0.004, 0.06) |  |
|  | 55 to <75 | 1.35 (0.75, 2.42) | 0.11 (0.06, 0.17) | 0.08 (0.06, 0.11) | 0.03 (-0.03, 0.09) |  |
|  | 75 plus | 0.76 (0.29, 2.03) | 0.11 (0.01, 0.21) | 0.15 (0.09, 0.20) | -0.03 (-0.15, 0.08) |  |
| Dermatology | <18 | 1.52 (0.65, 3.55) | 0.04 (0.01, 0.07) | 0.03 (0.02, 0.03) | 0.01 (-0.02, 0.04) | 0.96 |
|  | 18 to <35 | 1.31 (0.90, 1.91) | 0.06 (0.04, 0.07) | 0.04 (0.03, 0.05) | 0.01 (-0.01, 0.03) |  |
|  | 35 to <55 | 1.65 (1.01, 2.70) | 0.07 (0.04, 0.10) | 0.04 (0.03, 0.05) | 0.03 (-0.005, 0.06) |  |
|  | 55 to <75 | 1.50 (0.99, 2.28) | 0.12 (0.08, 0.16) | 0.08 (0.06, 0.10) | 0.04 (-0.005, 0.08) |  |
|  | 75 plus | 1.50 (0.91, 2.47) | 0.14 (0.07, 0.20) | 0.09 (0.07, 0.11) | 0.05 (-0.02, 0.11) |  |
| Medical Oncology | <18 | 51.9 (5.07, 531) | 0.01 (-0.01, 0.03) | <0.001 (<-0.001, <0.001) | 0.01 (-0.01, 0.03) | 0.007 |
|  | 18 to <35 | 5.07 (0.82, 31.1) | 0.002 (-0.001, 0.01) | <0.001 (<0.001, <0.001) | 0.002 (-0.002, 0.01) |  |
|  | 35 to <55 | 1.21 (0.51, 2.91) | 0.01 (0.003, 0.02) | 0.01 (0.01, 0.02) | 0.002 (-0.01, 0.01) |  |
|  | 55 to <75 | 1.72 (0.74, 3.98) | 0.04 (0.01, 0.06) | 0.02 (0.01, 0.03) | 0.02 (-0.01, 0.04) |  |
|  | 75 plus | 0.48 (0.14, 1.60) | 0.02 (-0.002, 0.05) | 0.05 (0.03, 0.08) | -0.03 (-0.07, 0.01) |  |
| Psychiatry | <18 | 2.91 (1.49, 5.68) | 0.07 (0.04, 0.10) | 0.02 (0.01, 0.04) | 0.05 (0.01, 0.08) | 0.06 |
|  | 18 to <35 | 6.25 (3.86, 10.1) | 0.14 (0.09, 0.19) | 0.02 (0.02, 0.03) | 0.12 (0.07, 0.17) |  |
|  | 35 to <55 | 5.33 (2.08, 13.7) | 0.25 (0.13, 0.38) | 0.05 (0.01, 0.08) | 0.20 (0.07, 0.33) |  |
|  | 55 to <75 | 3.34 (1.85, 6.03) | 0.13 (0.08, 0.18) | 0.04 (0.02, 0.06) | 0.09 (0.04, 0.15) |  |
|  | 75 plus | 1.90 (0.95, 3.80) | 0.15 (0.06, 0.25) | 0.08 (0.05, 0.11) | 0.07 (-0.03, 0.17) |  |
| Rehabilitation Medicine | <18 | 5.19 (0.75, 35.9) | 0.01 (-0.01, 0.03) | 0.002 (<0.001, 0.003) | 0.01 (-0.01, 0.02) | <0.001 |
|  | 18 to <35 | 3.12 (1.17, 8.33) | 0.01 (0.003, 0.01) | 0.003 (<0.001, 0.005) | 0.01 (<-0.001, 0.01) |  |
|  | 35 to <55 | 10.0 (4.14, 24.2) | 0.01 (0.01, 0.02) | 0.001 (<0.001, 0.002) | 0.01 (0.004, 0.02) |  |
|  | 55 to <75 | 0.70 (0.24, 2.07) | 0.01 (0.001, 0.02) | 0.02 (0.01, 0.03) | -0.005 (-0.02, 0.01) |  |
|  | 75 plus | 23.8 (4.30, 132) | 0.12 (-0.07, 0.31) | 0.01 (0.001, 0.01) | 0.12 (-0.07, 0.31) |  |
| Rheumatology | <18 | 4.51 (1.15, 17.7) | 0.01 (<0.001, 0.02) | 0.002 (<0.001, 0.005) | 0.01 (-0.003, 0.02) | 0.09 |
|  | 18 to <35 | 3.92 (1.99, 7.75) | 0.02 (0.01, 0.04) | 0.01 (0.004, 0.01) | 0.02 (0.004, 0.03) |  |
|  | 35 to <55 | 2.19 (1.39, 3.46) | 0.08 (0.05, 0.11) | 0.04 (0.03, 0.05) | 0.04 (0.01, 0.07) |  |
|  | 55 to <75 | 1.52 (0.94, 2.47) | 0.09 (0.05, 0.12) | 0.06 (0.04, 0.07) | 0.03 (-0.01, 0.07) |  |
|  | 75 plus | 1.25 (0.69, 2.26) | 0.06 (0.03, 0.09) | 0.05 (0.03, 0.07) | 0.01 (-0.02, 0.05) |  |
| Gynaecology | <18 | 0.91 (0.37, 2.21) | 0.03 (0.01, 0.06) | 0.04 (0.02, 0.05) | -0.003 (-0.03, 0.03) | 0.90 |
|  | 18 to <35 | 1.30 (1.04, 1.62) | 0.23 (0.19, 0.28) | 0.18 (0.16, 0.20) | 0.05 (0.004, 0.10) |  |
|  | 35 to <55 | 1.26 (1.00, 1.59) | 0.19 (0.15, 0.23) | 0.15 (0.14, 0.17) | 0.04 (-0.002, 0.08) |  |
|  | 55 to <75 | 1.26 (0.82, 1.93) | 0.10 (0.06, 0.13) | 0.08 (0.06, 0.09) | 0.02 (-0.02, 0.06) |  |
|  | 75 plus | 1.66 (0.78, 3.53) | 0.09 (0.03, 0.14) | 0.05 (0.03, 0.07) | 0.03 (-0.03, 0.10) |  |
| Non-clinical/hospital consultant led | <18 | 5.29 (2.68, 10.4) | 0.05 (0.02, 0.08) | 0.01 (0.01, 0.01) | 0.04 (0.01, 0.08) | 0.008 |
|  | 18 to <35 | 1.64 (1.12, 2.41) | 0.04 (0.02, 0.05) | 0.02 (0.02, 0.03) | 0.01 (0.001, 0.03) |  |
|  | 35 to <55 | 2.02 (1.47, 2.76) | 0.12 (0.09, 0.15) | 0.06 (0.05, 0.07) | 0.06 (0.03, 0.09) |  |
|  | 55 to <75 | 2.12 (1.47, 3.07) | 0.26 (0.18, 0.34) | 0.12 (0.10, 0.15) | 0.14 (0.05, 0.22) |  |
|  | 75 plus | 1.45 (0.86, 2.45) | 0.26 (0.14, 0.38) | 0.18 (0.13, 0.22) | 0.08 (-0.05, 0.21) |  |

**SA 5b: Admitted Patient Care**

| **Outcome** | **Age group** | **Rate Ratio (95% CI)** | **Rate per person-year in narcolepsy group  (95% CI)** | **Rate per person-year in comparison group  (95% CI)** | **Rate Difference per person-year  (95% CI)** | **Interaction p-value (95% CI)** |
| --- | --- | --- | --- | --- | --- | --- |
| Respiratory Medicine | <18 | 40.8 (12.2, 136) | 0.02 (0.01, 0.03) | <0.001 (<-0.001, <0.001) | 0.02 (0.01, 0.03) | <0.001 |
|  | 18 to <35 | 32.8 (21.1, 51.0) | 0.04 (0.04, 0.05) | 0.001 (<0.001, 0.002) | 0.04 (0.03, 0.05) |  |
|  | 35 to <55 | 27.6 (18.7, 40.9) | 0.06 (0.05, 0.07) | 0.002 (0.001, 0.003) | 0.06 (0.04, 0.07) |  |
|  | 55 to <75 | 8.34 (5.99, 11.6) | 0.05 (0.03, 0.06) | 0.01 (0.004, 0.01) | 0.04 (0.03, 0.05) |  |
|  | 75 plus | 3.93 (1.75, 8.85) | 0.05 (0.01, 0.09) | 0.01 (0.01, 0.02) | 0.04 (-0.001, 0.08) |  |
| Neurology | <18 | 139 (18.9, 1024) | 0.01 (0.01, 0.02) | <0.001 (<-0.001, <0.001) | 0.01 (0.01, 0.02) | <0.001 |
|  | 18 to <35 | 32.0 (17.7, 58.0) | 0.02 (0.02, 0.02) | <0.001 (<0.001, <0.001) | 0.02 (0.02, 0.02) |  |
|  | 35 to <55 | 18.2 (7.16, 46.3) | 0.03 (0.02, 0.04) | 0.002 (<0.001, 0.003) | 0.03 (0.02, 0.04) |  |
|  | 55 to <75 | 10.1 (5.93, 17.2) | 0.02 (0.01, 0.02) | 0.001 (<0.001, 0.002) | 0.01 (0.01, 0.02) |  |
|  | 75 plus | 5.78 (2.69, 12.4) | 0.01 (0.01, 0.02) | 0.002 (0.001, 0.003) | 0.01 (0.003, 0.02) |  |
| Ear Nose and Throat | <18 | 1.34 (0.83, 2.15) | 0.01 (0.01, 0.02) | 0.01 (0.01, 0.01) | 0.003 (-0.003, 0.01) | 0.02 |
|  | 18 to <35 | 2.52 (1.77, 3.58) | 0.01 (0.01, 0.02) | 0.01 (0.004, 0.01) | 0.01 (0.004, 0.01) |  |
|  | 35 to <55 | 3.77 (2.57, 5.54) | 0.02 (0.01, 0.02) | 0.004 (0.004, 0.01) | 0.01 (0.01, 0.02) |  |
|  | 55 to <75 | 2.76 (1.78, 4.29) | 0.01 (0.01, 0.02) | 0.01 (0.004, 0.01) | 0.01 (0.004, 0.01) |  |
|  | 75 plus | 1.82 (0.86, 3.86) | 0.01 (0.003, 0.01) | 0.004 (0.002, 0.01) | 0.003 (-0.002, 0.01) |  |
| General Surgery | <18 | 2.44 (1.48, 4.03) | 0.02 (0.01, 0.03) | 0.01 (0.01, 0.01) | 0.01 (0.004, 0.02) | 0.14 |
|  | 18 to <35 | 2.31 (1.80, 2.96) | 0.04 (0.03, 0.05) | 0.02 (0.01, 0.02) | 0.02 (0.01, 0.03) |  |
|  | 35 to <55 | 2.40 (1.97, 2.92) | 0.07 (0.06, 0.08) | 0.03 (0.03, 0.03) | 0.04 (0.03, 0.05) |  |
|  | 55 to <75 | 1.96 (1.66, 2.32) | 0.10 (0.08, 0.11) | 0.05 (0.05, 0.05) | 0.05 (0.03, 0.06) |  |
|  | 75 plus | 1.59 (1.19, 2.12) | 0.09 (0.07, 0.12) | 0.06 (0.05, 0.07) | 0.03 (0.01, 0.06) |  |
| Trauma and Orthopaedics | <18 | 1.78 (1.07, 2.95) | 0.02 (0.01, 0.02) | 0.01 (0.01, 0.01) | 0.01 (<-0.001, 0.01) | 0.08 |
|  | 18 to <35 | 1.34 (0.93, 1.95) | 0.01 (0.01, 0.02) | 0.01 (0.01, 0.01) | 0.003 (-0.001, 0.01) |  |
|  | 35 to <55 | 2.32 (1.81, 2.99) | 0.03 (0.03, 0.04) | 0.01 (0.01, 0.02) | 0.02 (0.01, 0.03) |  |
|  | 55 to <75 | 1.92 (1.49, 2.48) | 0.06 (0.04, 0.07) | 0.03 (0.03, 0.03) | 0.03 (0.01, 0.04) |  |
|  | 75 plus | 1.51 (1.15, 1.99) | 0.07 (0.05, 0.08) | 0.04 (0.04, 0.05) | 0.02 (0.01, 0.04) |  |
| Urology | <18 | 6.34 (2.09, 19.2) | 0.01 (<0.001, 0.02) | 0.002 (<0.001, 0.003) | 0.01 (-0.002, 0.02) | 0.07 |
|  | 18 to <35 | 3.44 (1.83, 6.48) | 0.02 (0.01, 0.02) | 0.005 (0.003, 0.01) | 0.01 (0.002, 0.02) |  |
|  | 35 to <55 | 3.42 (2.28, 5.14) | 0.02 (0.02, 0.03) | 0.01 (0.01, 0.01) | 0.02 (0.01, 0.03) |  |
|  | 55 to <75 | 1.61 (1.02, 2.54) | 0.04 (0.03, 0.06) | 0.03 (0.02, 0.03) | 0.02 (-0.002, 0.03) |  |
|  | 75 plus | 2.57 (1.36, 4.85) | 0.11 (0.04, 0.17) | 0.04 (0.03, 0.05) | 0.06 (0.002, 0.13) |  |
| Cardiology | <18 | 10.8 (2.98, 39.1) | 0.005 (<-0.001, 0.01) | <0.001 (<0.001, <0.001) | 0.005 (<-0.001, 0.01) | 0.02 |
|  | 18 to <35 | 3.58 (1.58, 8.12) | 0.004 (0.001, 0.01) | 0.001 (<0.001, 0.002) | 0.003 (<0.001, 0.01) |  |
|  | 35 to <55 | 2.66 (1.75, 4.03) | 0.01 (0.01, 0.02) | 0.005 (0.003, 0.01) | 0.01 (0.004, 0.01) |  |
|  | 55 to <75 | 2.57 (1.94, 3.42) | 0.04 (0.03, 0.06) | 0.02 (0.01, 0.02) | 0.03 (0.02, 0.04) |  |
|  | 75 plus | 1.47 (0.97, 2.24) | 0.05 (0.03, 0.06) | 0.03 (0.03, 0.04) | 0.01 (-0.004, 0.03) |  |
| Endocrinology and Diabetes | <18 | 31.5 (7.28, 136) | 0.01 (<0.001, 0.01) | <0.001 (<-0.001, <0.001) | 0.01 (<0.001, 0.01) | <0.001 |
|  | 18 to <35 | 5.91 (2.38, 14.6) | 0.004 (0.001, 0.01) | <0.001 (<0.001, 0.001) | 0.003 (<0.001, 0.01) |  |
|  | 35 to <55 | 6.07 (3.22, 11.4) | 0.01 (0.003, 0.01) | 0.001 (<0.001, 0.001) | 0.01 (0.002, 0.01) |  |
|  | 55 to <75 | 8.36 (4.82, 14.5) | 0.01 (0.01, 0.02) | 0.001 (<0.001, 0.002) | 0.01 (0.01, 0.02) |  |
|  | 75 plus | 1.45 (0.72, 2.92) | 0.01 (0.003, 0.01) | 0.01 (0.004, 0.01) | 0.003 (-0.003, 0.01) |  |
| General Internal Medicine | <18 | 3.66 (1.53, 8.79) | 0.01 (0.002, 0.01) | 0.001 (<0.001, 0.002) | 0.004 (<0.001, 0.01) | 0.02 |
|  | 18 to <35 | 3.07 (2.27, 4.16) | 0.05 (0.04, 0.06) | 0.02 (0.01, 0.02) | 0.03 (0.02, 0.05) |  |
|  | 35 to <55 | 3.93 (3.14, 4.91) | 0.10 (0.08, 0.12) | 0.03 (0.02, 0.03) | 0.08 (0.06, 0.09) |  |
|  | 55 to <75 | 3.12 (2.44, 4.00) | 0.19 (0.16, 0.23) | 0.06 (0.05, 0.07) | 0.13 (0.09, 0.17) |  |
|  | 75 plus | 2.31 (1.87, 2.84) | 0.32 (0.26, 0.37) | 0.14 (0.12, 0.15) | 0.18 (0.12, 0.24) |  |
| Clinical Haematology | <18 | 3.74 (0.37, 37.8) | 0.01 (-0.003, 0.01) | 0.002 (-0.001, 0.004) | 0.004 (-0.01, 0.01) | 0.40 |
|  | 18 to <35 | 1.51 (0.17, 13.3) | 0.01 (-0.01, 0.04) | 0.01 (-0.001, 0.02) | 0.004 (-0.02, 0.03) |  |
|  | 35 to <55 | 0.94 (0.27, 3.22) | 0.01 (<-0.001, 0.01) | 0.01 (0.003, 0.01) | <-0.001 (-0.01, 0.01) |  |
|  | 55 to <75 | 2.46 (0.78, 7.73) | 0.03 (<0.001, 0.07) | 0.01 (0.01, 0.02) | 0.02 (-0.01, 0.05) |  |
|  | 75 plus | 0.49 (0.11, 2.26) | 0.01 (-0.002, 0.02) | 0.02 (0.002, 0.04) | -0.01 (-0.03, 0.01) |  |
| Psychiatry | <18 | 4.57 (0.38, 54.3) | 0.01 (-0.01, 0.02) | 0.002 (-0.001, 0.01) | 0.01 (-0.01, 0.02) | 0.34 |
|  | 18 to <35 | 3.76 (1.32, 10.7) | 0.01 (0.003, 0.02) | 0.004 (<0.001, 0.01) | 0.01 (<-0.001, 0.02) |  |
|  | 35 to <55 | 6.76 (3.90, 11.7) | 0.01 (0.01, 0.01) | 0.002 (<0.001, 0.002) | 0.01 (0.01, 0.01) |  |
|  | 55 to <75 | 4.60 (1.67, 12.7) | 0.01 (0.001, 0.02) | 0.002 (<0.001, 0.003) | 0.01 (<-0.001, 0.01) |  |
|  | 75 plus | 2.79 (1.24, 6.26) | 0.01 (0.002, 0.01) | 0.003 (0.002, 0.004) | 0.01 (<-0.001, 0.01) |  |
| Non-clinical/hospital consultant led | <18 | 116 (14.0, 962) | 0.05 (-0.04, 0.13) | <0.001 (<-0.001, <0.001) | 0.05 (-0.04, 0.13) | <0.001 |
|  | 18 to <35 | 2.60 (0.62, 11.0) | 0.004 (-0.001, 0.01) | 0.002 (<0.001, 0.003) | 0.003 (-0.003, 0.01) |  |
|  | 35 to <55 | 1.00 (0.51, 1.96) | 0.01 (0.004, 0.01) | 0.01 (0.01, 0.01) | <0.001 (-0.01, 0.01) |  |
|  | 55 to <75 | 1.41 (0.70, 2.81) | 0.02 (0.01, 0.03) | 0.01 (0.01, 0.02) | 0.01 (-0.01, 0.02) |  |
|  | 75 plus | 1.16 (0.48, 2.81) | 0.02 (0.01, 0.03) | 0.01 (0.01, 0.02) | 0.002 (-0.01, 0.02) |  |

**SA Table 6 Rate Ratios comparing the rate of events in each setting in the five-years before and after diagnosis for people with narcolepsy and the comparison group using Poisson and Negative binomial regression models**

| **NHS Setting/ Regression model** | **Rate ratio (95% CI)** | **Dispersion parameter (95% CI)** |
| --- | --- | --- |
| **Outpatient** |  |  |
| Poisson model (primary) | 2.73 (2.58, 2.89) |  |
| Negative binomial regression | 3.84 (3.51, 4.20) | 1.24 |
| **Admitted Patient Care** |  |  |
| Poisson model (primary) | 2.45 (2.14, 2.82) |  |
| Negative binomial regression | model did not converge |  |
| **Accident & Emergency** |  |  |
| Poisson model (primary) | 2.21 (2.05, 2.39) |  |
| Negative binomial regression | model did not converge |  |
| **Primary care** |  |  |
| Poisson model (primary) | 1.85 (1.77, 1.93) |  |
| Negative binomial regression | 2.06 (1.95, 2.19) | 0.91 |

**SA Table 7: Rate Ratios and Rate Differences comparing the rate of events to each NHS setting in the five-years before and after diagnosis for people with narcolepsy and the comparison group using alternative narcolepsy definitions**

| **Outcome** | **Narcolepsy Definition** | **Rate ratio (95% CI)** | **Rate Difference per person-year (95% CI)** |
| --- | --- | --- | --- |
| **Outpatient** |  |  |  |
| All events | Primary care or HES APC | 2.73 (2.58, 2.89) | 1.91 (1.76, 2.05) |
|  | Primary care only | 2.62 (2.46, 2.80) | 1.62 (1.48, 1.76) |
| Possible sleep-related events | Primary care or HES APC | 9.01 (8.30, 9.79) | 0.77 (0.73, 0.81) |
|  | Primary care only | 9.54 (8.66, 10.5) | 0.78 (0.73, 0.83) |
| **Admitted Patient Care** |  |  |  |
| All events | Primary care or HES APC | 2.45 (2.14, 2.82) | 0.36 (0.30, 0.42) |
|  | Primary care only | 2.09 (1.86, 2.35) | 0.24 (0.20, 0.28) |
| Possible sleep-related events | Primary care or HES APC | 7.81 (6.49, 9.40) | 0.10 (0.09, 0.11) |
|  | Primary care only | 7.71 (6.20, 9.58) | 0.09 (0.08, 0.10) |
| **Accident & Emergency** |  |  |  |
| All events | Primary care or HES APC | 2.21 (2.05, 2.39) | 0.25 (0.22, 0.29) |
|  | Primary care only | 1.83 (1.68, 2.00) | 0.17 (0.14, 0.20) |
| **Primary care** |  |  |  |
| All events | Primary care or HES APC | 1.85 (1.77, 1.93) | 4.32 (3.95, 4.69) |
|  | Primary care only | 1.76 (1.68, 1.84) | 3.59 (3.24, 3.94) |

**SA Figure 1: Rates of healthcare resource use in the narcolepsy and comparison groups over time relative to the index date**

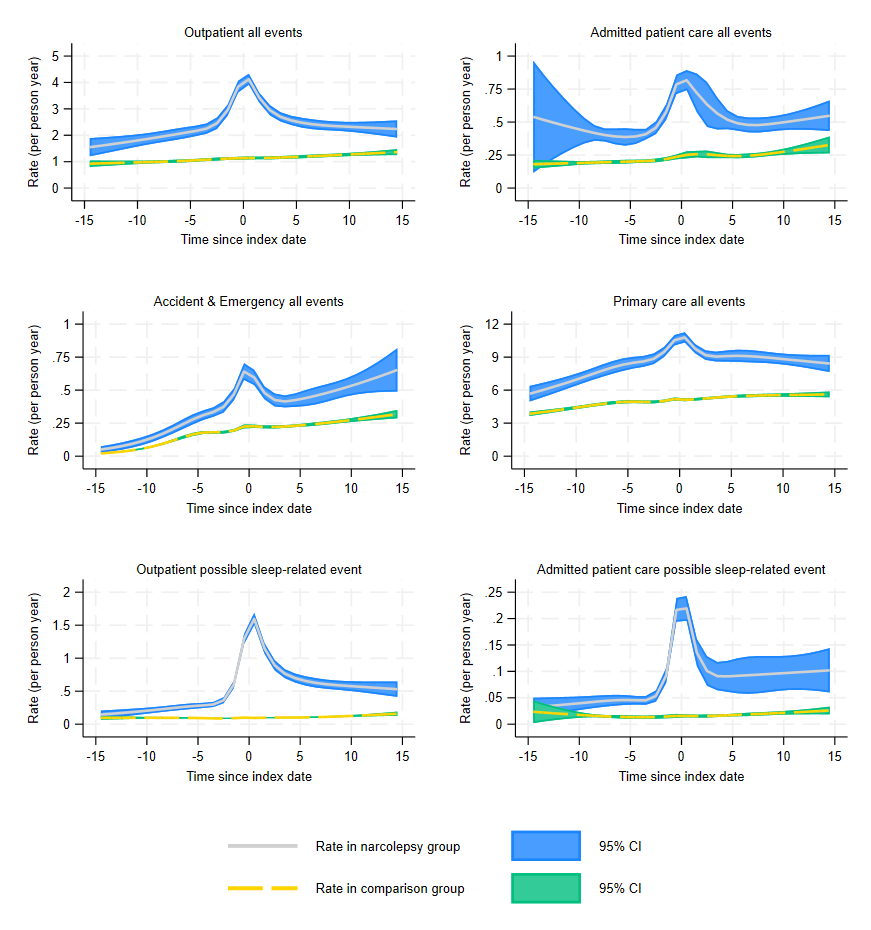

**SA Figure 2: Rate Ratios comparing healthcare resource use in the narcolepsy and comparison groups over time relative to the index date**

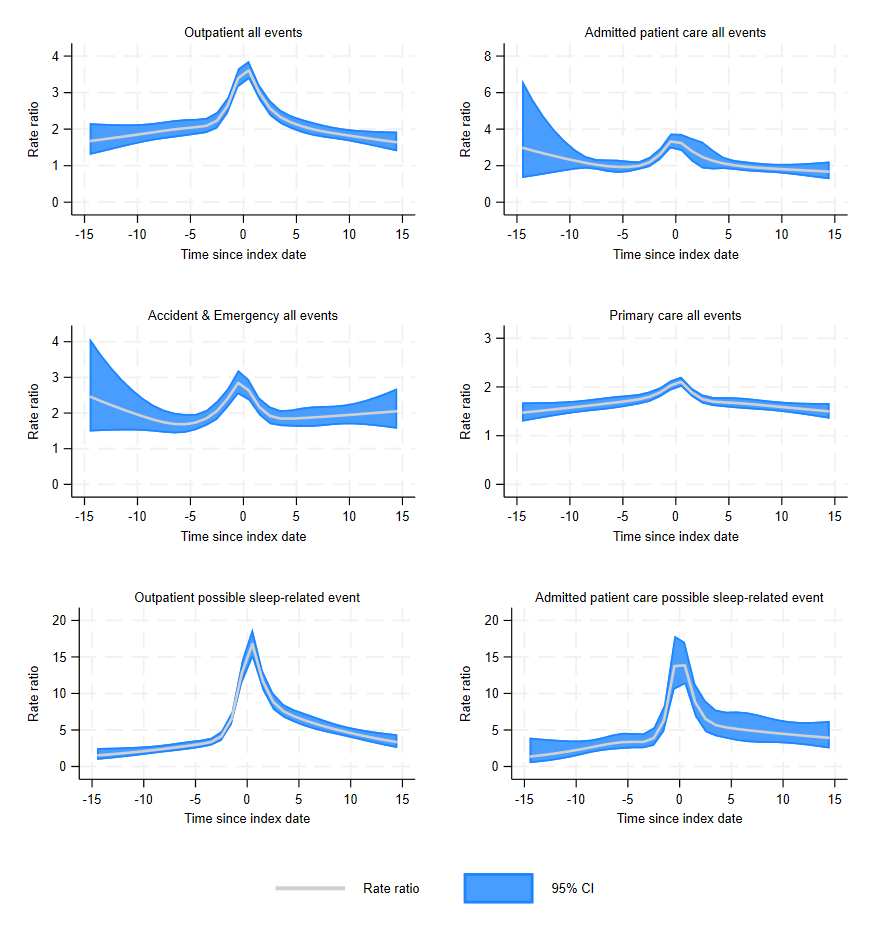
